# Gene discovery and pleiotropic architecture of Chronic Pain in a Genome-wide Association Study of >1.2 million Individuals

**DOI:** 10.1101/2025.02.28.25323112

**Authors:** Sylvanus Toikumo, Christal Davis, Zeal Jinwala, Yousef Khan, Mariela Jennings, Lea Davis, Sandra Sanchez-Roige, Rachel L. Kember, Henry R. Kranzler

## Abstract

Chronic pain is highly prevalent worldwide, and genome-wide association studies (GWAS) have identified a growing number of chronic pain loci. To further elucidate its genetic architecture, we leveraged data from 1,235,695 European ancestry individuals across three biobanks. In a meta-analytic GWAS, we identified 343 independent loci for chronic pain, 92 of which were new. Sex-specific meta-analyses revealed 115 independent loci (12 of which were new) for males (N = 583,066) and 12 loci (two of which were new) for females (N = 241,266). Multi-omics gene prioritization analyses highlighted 490 genes associated with chronic pain through their effects on brain- and blood-specific regulation. Loci associated with increased risk for chronic pain were also associated with increased risk for multiple other traits, with Mendelian randomization analyses showing that chronic pain was causally associated with psychiatric disorders, substance use disorders, and C-reactive protein levels. Chronic pain variants also exhibited pleiotropic associations with cortical area brain structures. This study expands our knowledge of the genetics of chronic pain and its pathogenesis, highlighting the importance of its pleiotropy with multiple disorders and elucidating its multi-omic pathophysiology.

## INTRODUCTION

Chronic pain, which is disproportionately prevalent among women, is associated with high rates of morbidity and mortality and a high socio-economic toll on affected individuals and their families^1^. Despite its substantial and growing public health impact, there are no clinically useful objective indicators of chronic pain^2^, which if available, could enhance preventive and therapeutic interventions to address it. Genomics, including genome-wide association studies (GWAS), shows promise for identifying biomarkers for chronic pain.

To date, GWASs have identified over 170 risk loci for chronic pain^3–5^. Despite their diversity, pain conditions are highly overlapping^6^, sharing a common genetic risk profile^7,8^ and similar alterations in the central nervous system^9,10^. Structural and functional changes in the brain are thought to reflect the transition from acute to chronic pain^11,12^. Substantiating these findings using genomics could provide diagnostic and predictive tools to advance personalized pain medicine^13,14^ and support the development of mechanism-specific pain treatments^15,16^. Furthermore, although chronic pain often co-occurs with psychiatric (e.g., major depression), cardiovascular (e.g., hypertension), and metabolic (e.g., diabetes) disorders ^5,17^, the nature of these associations, including whether they are causal, is unknown.

To address these important questions, we conducted a meta-analysis of three independent GWAS of chronic pain (*N* = 1,235,695), followed by a sex-specific meta-analysis in a subsample (male, *N* = 583,066 and female, *N* = 241,266) (Figure 1). We then integrated multiple layers of functional omics data to identify effector genes for chronic pain. To refine our understanding of risk factors for chronic pain and clarify the relations among them, we conducted extensive genetic correlation, univariate and bivariate causal mixture models (using MiXeR), and Mendelian randomization (MR) analyses, a phenome-wide association study (PheWAS) in two independent biobanks and lab-wide association study (LabWAS).

**Figure 1.**
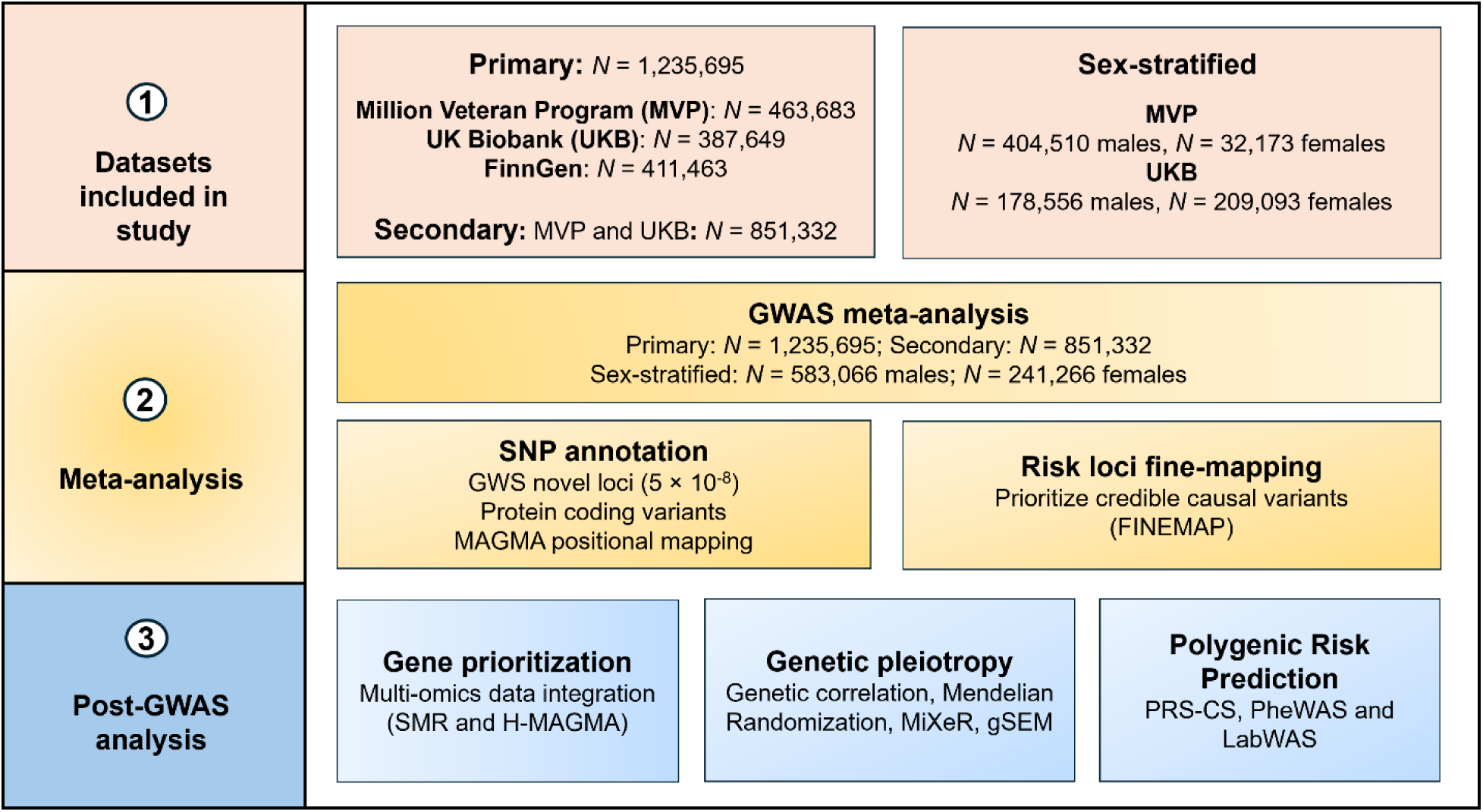
Overview of the study.

## RESULTS

### Genome-wide association analysis

The meta-analysis comprised summary statistics from prior published GWAS of pain conducted in three large biobanks (Supplementary Table 1)^3,5,18^. Genetic correlations, *r*_g_, for the traits across biobanks were moderately high and positive (Million Veteran Program (MVP) vs UK Biobank (UKB): *r*_g_ > 0.79, *P* < 2.23 × 10^−308^, MVP vs FinnGen: *r*_g_ = 0.66, *P* = 1.30 × 10^−163^, and UKB vs FinnGen: *r*_g_ = 0.69, *P* < 2.97 × 10^−213^; Figure 2A and Supplementary Table 2), reflecting the similarity of the genetic architecture of the pain phenotypes across biobanks and supporting the utility of meta-analysis.

**Figure 2.**
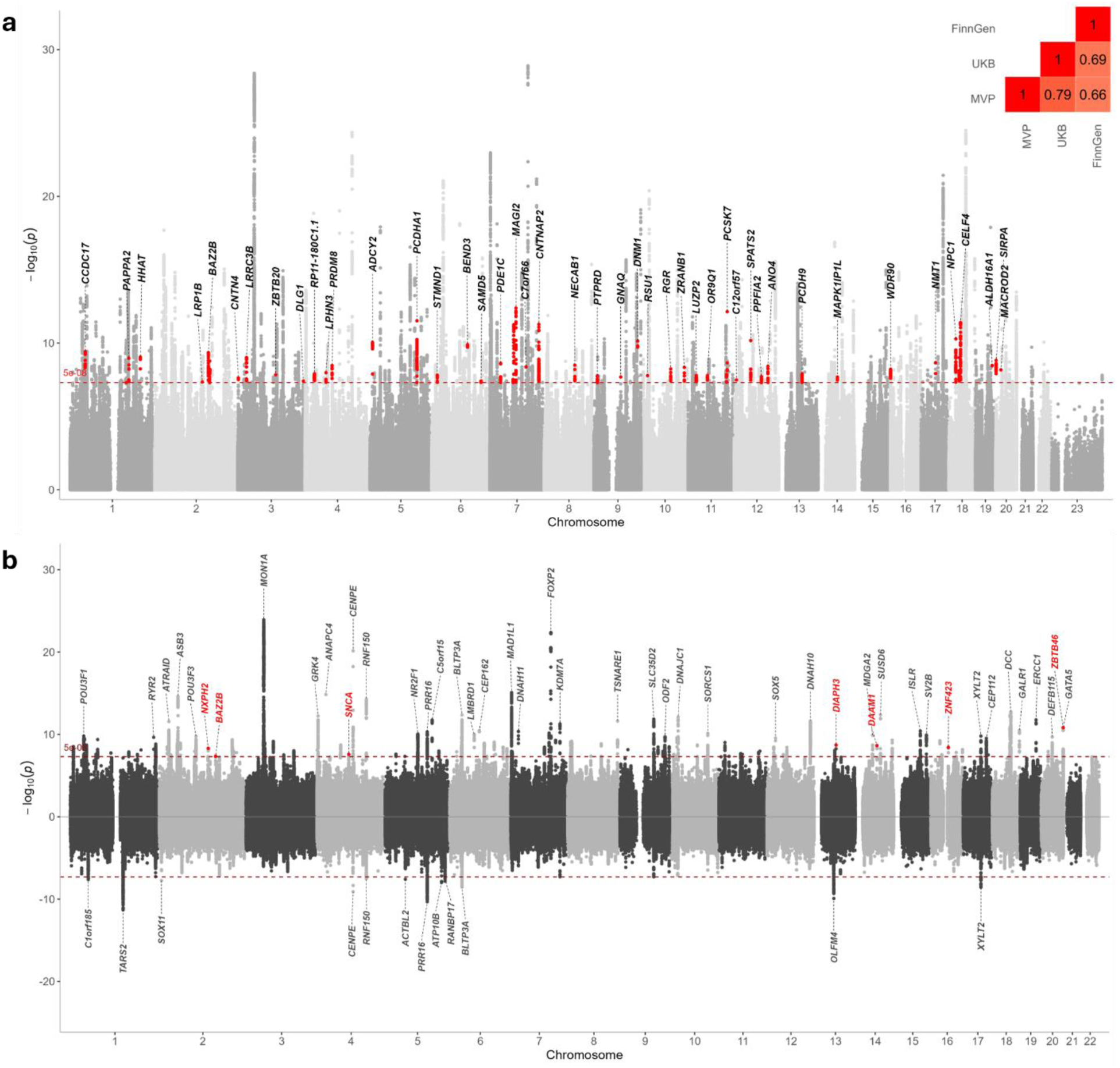
**a**) Chronic pain meta-analysis identified 343 independent risk loci including 92 new pain associations, of which 42 (highlighted in bold) are protein-coding genes **b**) Sex-specific meta-analyses of pain identified 115 independent loci in males (top), including 12 new association signals, of which 7 are protein-coding (highlighted in red) and 12 loci in females (bottom).

In our meta-analysis, genomic inflation factors (*λ*_GC_) values were 1.68, 1.49, 1.33, and 1.21 for the primary, secondary, male, and female GWAS, respectively (Extended Data Figure 1). The intercepts (primary: 1.2, s.e. = 0.02, secondary: 1.1, s.e. = 0.01, male: 1.1, s.e. = 0.01, female: 1.0, s.e. = 0.01) and attenuation ratios (primary: 0.09, s.e. = 0.01, secondary: 0.08, s.e. = 0.01, male: 0.09, s.e. = 0.01, female: 0.03, s.e. = 0.02) (Supplementary Table 3) of the LDSC regression^19^ suggest that the elevated lambdas and inflation in the quantile plot are due to polygenicity rather than population stratification.

The primary meta-analysis identified 20,242 genome-wide significant (GWS) associations, which we resolved to 343 statistically independent signals using conditional and joint (COJO) SNP analysis (Methods), which accounts for linkage disequilibrium (LD) among nearby variants (Supplementary Table 4). Of the implicated loci, 92 were new association signals for chronic pain and 42 were protein-coding variants (Figure 2A).

In the secondary meta-analysis that comprised MVP and UKB only, we identified 11,767 SNPs in 212 independent risk loci (Supplementary Table 5). Of these, most loci (n=154 or 72.6%) were linked to chronic pain in the primary meta-analysis, 55 (25.9%) loci were not GWS in the primary analysis, and 3 SNPs (1.4%) were not present at all in the primary meta-analysis. Of the loci limited to the secondary meta-analysis, 14 have not previously been associated with pain (e.g., *DLG1*-rs11927266, *TNRC18-*rs12532973, *PIH1D1*-rs145838789, and *PTPRT*-rs1006749) (Supplementary Table 5).

SNP heritability (*h^2^*) was modest for chronic pain overall (primary, *h^2^* = 0.05 and secondary, *h^2^* = 0.07) and separately by sex (females, (*h^2^* = 0.08) and males, *h^2^* = 0.07) (Supplementary Table 3). The difference in *h*^2^ between the primary and secondary meta-analyses is likely due to the exclusion of the FinnGen sample, in which there is a founder effect^18^ and for which pain was defined broadly as a composite phenotype using one or more documented International Classification of Diseases – 9^th^ and 10^th^ Revision (ICD-9 and -10) diagnostic codes for 16 pain disorders (Supplementary Table 1). Because the primary meta-analysis yielded more risk loci, we based all downstream analyses (except *r*_g_ analyses) on the GWAS results from that sample.

Using a minimum physical distance of 1 MB (+/-) from the LD block of the lead SNP, we identified 215 genomic regions for the 343 independent loci (Supplementary Table 6) and fine-mapped the regions using the FINEMAP Bayesian method (Methods). For regions with independent causal signals (Supplementary Table 6), credible sets of variants were constructed to capture 95% of the regional posterior probability (Supplementary Table 7). Among the 343 independent loci, 68 were fine-mapped to a 95% credible set, of which 46 harbored known protein-coding genes and 16 were new chronic pain associations (Posterior inclusion probability, PP>0.99: *ANO4-*rs76904423, *NMT1-*rs12936234, *ALDH16A1-*rs10401643, *MACROD2-*rs80044408, *DLG1-* rs145646459, and *ZBTB20-*rs9837485; Supplementary Table 7).

To detect potential sex differences in the genetic influences on chronic pain, we performed meta-analyses separately for males and females using GWAS data from MVP^5^ and UKB^20^; similar data were not available for FinnGen. Genetic correlation was high and positive across biobanks (males, MVP vs UKB: *r*_g_ = 0.79, *P* = 4.36 × 10^−164^, and females, UKB vs MVP: *r*_g_ = 0.70, *P* < 3.62 × 10^−22^) and sexes (*r*_g_ > 0.88, *P* < 3.90 × 10^−281^) (Supplementary Table 2).

Among males, 5,526 SNPs at 115 independent loci were associated with chronic pain, with 26 observed only in males and 12 (including 7 protein-coding genes) not previously reported in pain GWAS (Figure 2B, Supplementary Table 8). Among females, there were 475 GWS SNPs at 12 independent loci (Figure 2B), of which two (*MAML3*-rs7675956 and *GABRB2*-rs1946247) were not GWS in the input GWASs and males (Supplementary Table 9). The direction of allelic effects was highly correlated between males and females (*r* = 0.71, *P* = 2.2 × 10^−16^; Extended Data Figure 2).

### Gene prioritization

By integrating GWAS signals with gene-expression data, we found significant colocalization (HEIDI *p* > 0.05) at 66 loci (Supplementary Table 10), representing 43 potentially causal genes. Among the 42 new chronic pain loci that harbor protein-coding variants (Supplementary Tables 4), five genes (*HECTD3* (chr1.p34.3), *CCDC17* (chr1.p34.3), *DNM1* (chr9.q33.1), *PCDHA1* (chr5.q31.5), and *WDR90* (chr16.p13.3)) showed evidence of colocalization in more than one brain tissue. Additionally, we found 49 splicing quantitative trait loci (sQTLs) and 116 unique methylation trait quantitative loci (mQTLs) (53 brain and 77 blood mQTLs), of which 4 and 10, respectively, reside within new pain loci, including *CCDC17*, *DNM1*, *PCDHA1*, and *WDR90* (Supplementary Table 11 & 12). In total, 44 unique variants (22 brain and 27 blood pQTLs) showed significant cis associations with protein levels, of which four (ECM1, HINT1, LGALS3, and SIRPA) had concordant effects in brain and blood (Figure 3A, Supplementary Table 13 & 14). Two of the four protein quantitative trait loci (pQTLs) were localized to genomic regions – chr5.q31.5 (*HINT1*) and chr20.p13 (*SIRPA*) – that harbor new chronic pain loci (Supplementary Table 4). Using the chromatin-based approach, H-MAGMA, we identified 313 genes associated with all six chromatin annotations (Supplementary Table 15), of which 11 overlapped with other omics findings, including *CCDC17*, *DNM1, PCDHA1*, and *WDR90*.

**Figure 3.**
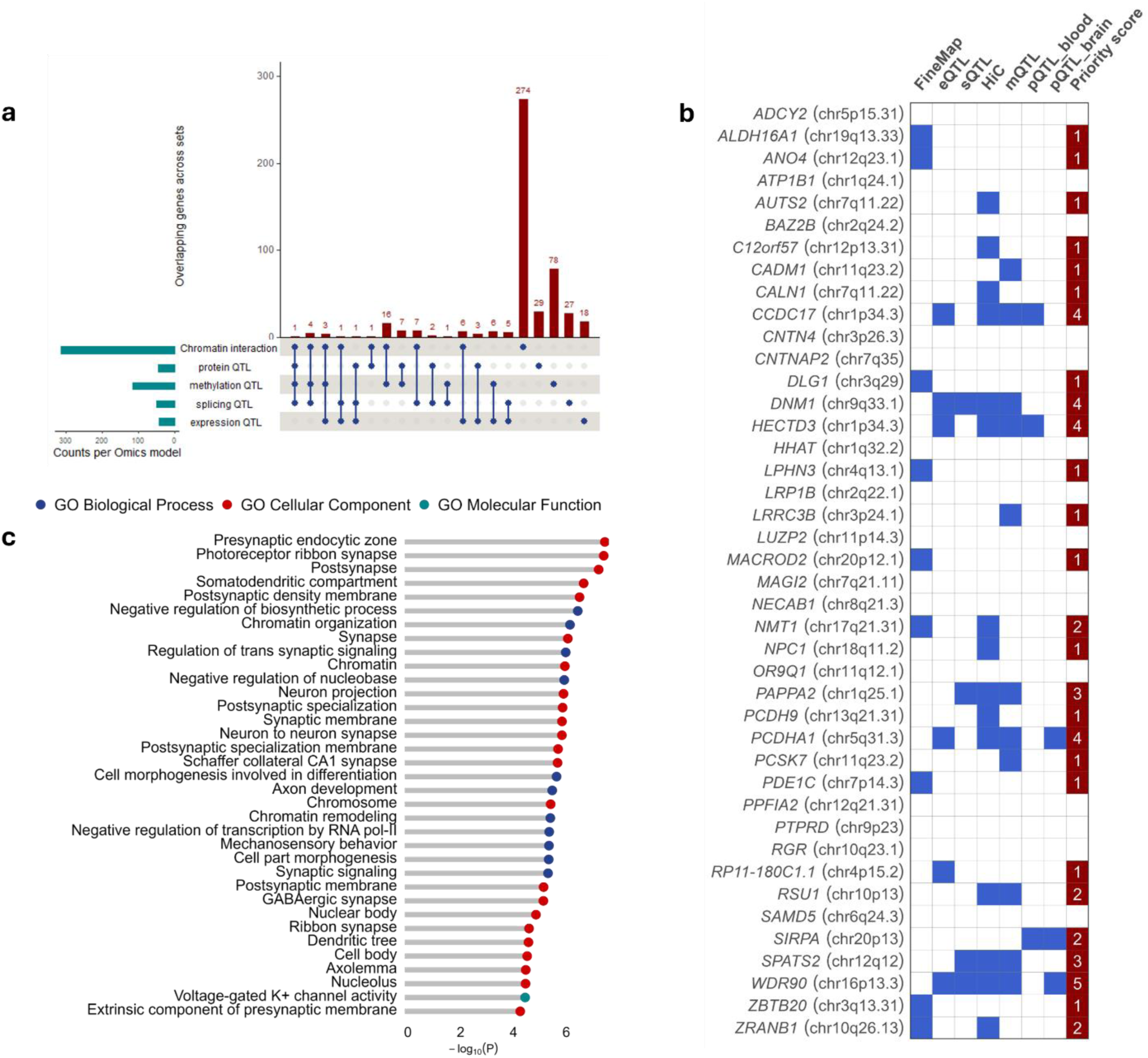
**a**) Gene prioritization for chronic pain using a multi-omics approach identified 490 genes, of which 64 overlap in at least two omics datasets **b**) Novel protein-coding GWAS loci for chronic pain (n = 42) that harbor genes prioritized from the multi-omics analysis. 11 of the 42 protein-coding new pain loci were supported by at least two lines of evidence **c**) MAGMA gene set enrichment analysis showing significantly enriched pathways for chronic pain (Bonferroni *P* = 4.75 × 10^−6^; one-sided t-test).

After controlling for multiple comparisons, we identified 490 unique genes with statistical evidence of association with chronic pain (Figure 3A, Supplementary Table 16). Seven of the 42 protein-coding new chronic pain loci were supported by at least three lines of evidence, and 11 by at least two lines of evidence (Figure 3B, Supplementary Table 17). Using MAGMA, GWAS variants successfully mapped to a nearby gene were enriched in gene pathways related to synaptic function, somatodendritic compartment, neuronal regulation and development, and axon development (Figure 3C).

### Evaluating a factor model of pain

We identified shared genetic risk between pain intensity and 29 heritable chronic pain conditions from the UKB (Extended Data Figure 3A). The confirmatory factor analyses (CFA) in which pain intensity loaded onto both the general and musculoskeletal pain factors (Extended Data Figure 3B) showed a significantly better fit than the model with pain intensity loading solely on the general pain factor (Δχ^2^(1) = 351.31, *P* < 0.001) (Extended Data Figure 3C). Additionally, the model in which pain intensity loaded onto both factors had slightly better Akaike information criterion (AIC) (7251.18 vs. 7600.49), comparative fit index (CFI) (0.944 vs. 0.941), and standardized root mean squared residual (SRMR) values (0.074 vs. 0.076) than the model without pain intensity included on the musculoskeletal factor. The general pain factor accounted for 57.14% and the musculoskeletal pain factor only 6.80% of the genetic variance in pain intensity.

### Genetic correlations with complex disease traits and brain structure

We estimated genetic correlations between chronic pain and 80 complex human phenotypes (Figure 4 and Supplementary Table 18). After Bonferroni correction, chronic pain was genetically correlated with 59 traits, including positive associations with 11 psychiatric (e.g., attention deficit hyperactivity disorder (ADHD), bipolar disorder, major depressive disorder (MDD), neuroticism, posttraumatic stress disorder (PTSD), PTSD symptoms, and Tourette Syndrome) and 10 substance use-related phenotypes (e.g., alcohol use disorder, cannabis use disorder, opioid use disorder, smoking initiation, substance use disorder (SUD) latent factor, and tobacco use disorder (TUD)). Chronic pain was also positively genetically correlated with levels of the immune biomarker C-reactive protein (CRP) and with insomnia and anthropometric or metabolic traits, including body mass index (BMI), obesity, Type 2 diabetes, and hip and waist circumference. Several medication-use phenotypes, including drug use classes N02A (opioids), A02B (peptic ulcer), N06A (antidepressants), N02BA (salicylic acid and derivatives), N02BE (anilides), N02C (antimigraine preparations), R03A (adrenergic inhalants), C01D (vasodilators used in cardiac disease), also showed positive genetic correlations with chronic pain. The genetic correlation patterns were similar between the primary and secondary meta-analyses, with two exceptions: the significant genetic correlation between chronic pain and bipolar disorder in the primary analysis was not present in the secondary analysis and the significant genetic correlation between pain and obsessive-compulsive disorder present in the secondary analysis was not seen in the primary one.

**Figure 4.**
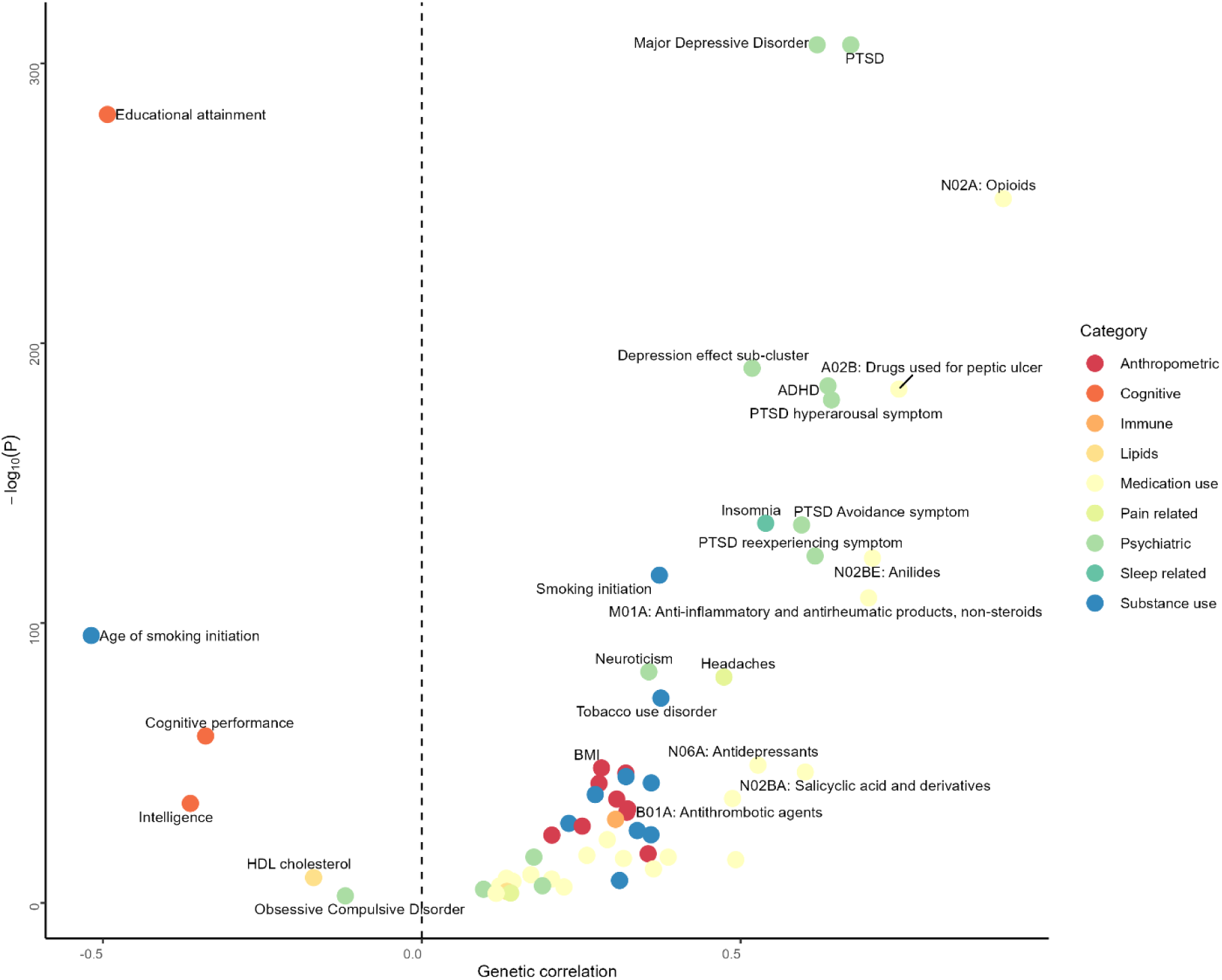
Genetic correlation for chronic pain using linkage disequilibrium score regression (LDSC). All traits passing Bonferroni correction (*P* = 6.25 × 10^−4^ (0.05/80)) are plotted. The color of the circle indicates the phenotypic category. The vertical dashed line represents genetic correlation = 0. BMI, body mass index; HDL, high-density lipoprotein; PTSD, posttraumatic stress disorder; ADHD, attention deficit/hypersensitivity disorder.

After Bonferroni correction, sex-specific analyses showed that chronic pain was significantly genetically correlated with 59 and 57 complex human phenotypes in females and males, respectively (Supplementary Table 18). Of these, five phenotypes (females only: bipolar disorder, migraine, PGC cross-disorder phenotype, and autism spectrum disorder; males only: obsessive-compulsive disorder) showed sex-specific genetic correlations with chronic pain.

We also examined genetic correlations between chronic pain and 1,323 brain image-derived phenotypes (IDPs) with significant heritability estimates^21^. After Bonferroni correction, 40 brain IDPs were genetically correlated with chronic pain (Supplementary Table 19). Among them, cortical thickness IDPs were positively genetically correlated with chronic pain: mean thickness of rostralmiddlefrontal (*r_g_* = 0.13; *P* = 8.58 × 10^−6^), S-front-middle (*r_g_*= 0.16; *P* = 1.85 × 10^−5^), and G-parietal-superior (*r_g_* = 0.12; *P* = 3.38 × 10^−5^) in the left hemisphere. Chronic pain was negatively genetically correlated with cortical area IDPs including the area of middletemporal in the left (*r_g_* = -0.16; *P* = 5.97 × 10^−10^) and right (*r_g_* = -0.15; *P* = 4.82 × 10^−9^) hemispheres and total surface area in the right hemisphere (*r_g_* = - 0.15; *P* = 1.81 × 10^−8^).

### Potential causal genetic effects

We used MR to assess potential causal genetic effects between chronic pain and the complex human traits (*n* = 59) and brain structures (*n* = 40) that displayed a significant genetic correlation after Bonferroni correction. We observed putative bidirectional causal genetic effects for seven psychiatric disorders (MDD, neuroticism, PTSD, PTSD avoidance and hyperarousal symptoms, and depression and worry sub-clusters), smoking initiation, and waist circumference (Figure 5A, Supplementary Table 20). We also found evidence that chronic pain was positively causally associated with SUDs (alcohol, cannabis, opioids, and tobacco), ADHD, insomnia, and CRP level (Figure 5A, Supplementary Table 20).

**Figure 5.**
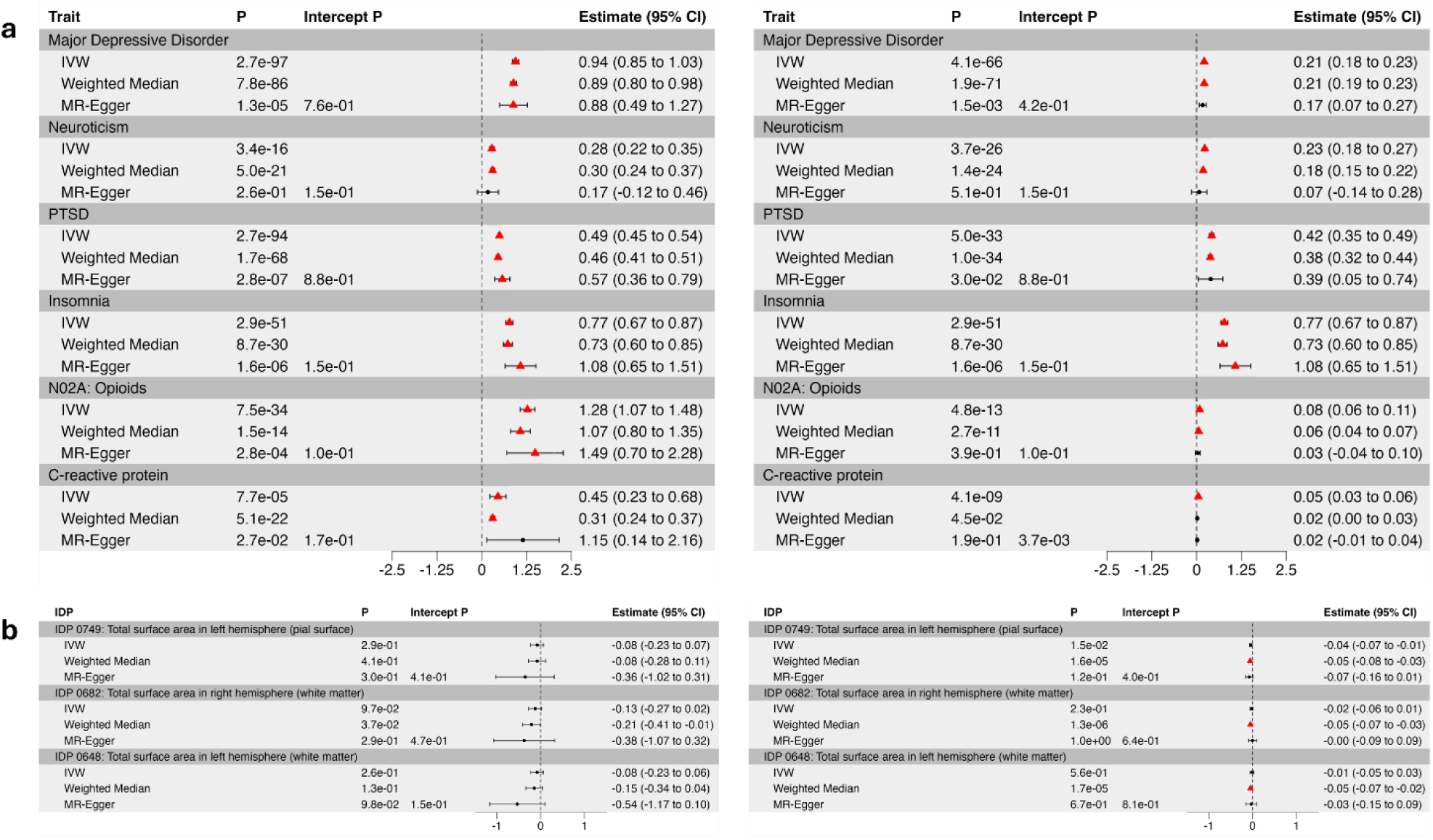
Causal association for (**a**) complex traits and (**b**) brain image-derived phenotypes (IDPs) genetically correlated with chronic pain. Left: Chronic pain as the exposure, traits/IDPs as the outcome. Right: Traits/IDPs as the exposure, chronic pain as the outcome. Estimates (+/- 95% CI) and p-values for each MR analysis [Inverse variance weighted (IVW), Weighted median, and MR-Egger] are shown. MR results that pass multiple tests (complex traits, *P* < 8.47×10^-4^; brain IDPs, *P* < 1.25 × 10^-3^) are indicated in red. Intercept p-value: MR-Egger horizontal pleiotropy test. Error bars: estimate +/- 95% CI.

Among medication use phenotypes, evidence for bidirectional causation was observed for N02A (opioids), A02B (peptic ulcer), N02BE (anilides), and N02C (antimigraine preparations) (Supplementary Table 20). Chronic pain was positively causally associated with N06A (antidepressants), N02BA (salicylic acid and derivatives), R03A (adrenergic inhalants), and C01D (vasodilators used in cardiac disease).

We identified significant (*P* < 0.05/40 = 1.25 × 10^−3^) negative putative causal effects of three cortical area IDPs on chronic pain (Figure 5B, Supplementary Table 21): total surface area in the left hemisphere based on parcellation of the white matter (Weighted mean, OR = 0.96, 95% CI of -0.07 to -0.03, *P* = 1.66 × 10^−5^), total surface area of the right hemisphere based on parcellation of the pial (Weighted mean, OR = 0.96, 95% CI of -0.07 to -0.03, *P* = 1.55 × 10^−5^), and white matter (Weighted mean, OR = 0.95, 95% CI of -0.07 to -0.03, *P* = 1.31 × 10^−6^). These findings are consistent with the observation that lower cortical surface area activity plays a role in chronic pain pathology^12^.

### Polygenic pleiotropic traits for chronic pain

For all traits, the univariate MiXeR models yielded positive AIC values, indicating that there was sufficient power to perform bivariate models. Chronic pain was estimated to have 11,262 (SD = 262.13) causal variants. Across the traits examined, we observed varying degrees of genetic overlap with chronic pain, generally exceeding the overlap predicted by the genetic correlation alone (reflected in positive AIC values for the MiXeR model compared to the minimum overlap model).

Insomnia, MDD, PTSD, and neuroticism shared a large proportion of causal variants with chronic pain, ranging from 81.26% (SD = 0.03) for the SUD latent factor to 93.09% (SD = 0.029) for neuroticism (Figure 6). Despite the consistently high overlap, the proportion of variants with concordant effects differed, with neuroticism showing a lower proportion (63.78%, SD = 0.004) than MDD (78.77%, SD = 0.075), PTSD (78.74%; SD = 0.033), and insomnia (75.29%, SD = 0.063). SUDs and opioid medication use showed a high degree of overlap with chronic pain. Opioid medication use shared 91.37% (SD = 0.030) of causal variants with chronic pain, nearly all (96.29%, SD = 0.039) of which had a concordant effect direction. The SUD factor also showed substantial overlap with chronic pain (81.26%, SD = 0.03), though only 63.60% (SD = 0.007) of shared variants were concordant in direction across the two traits.

**Figure 6.**
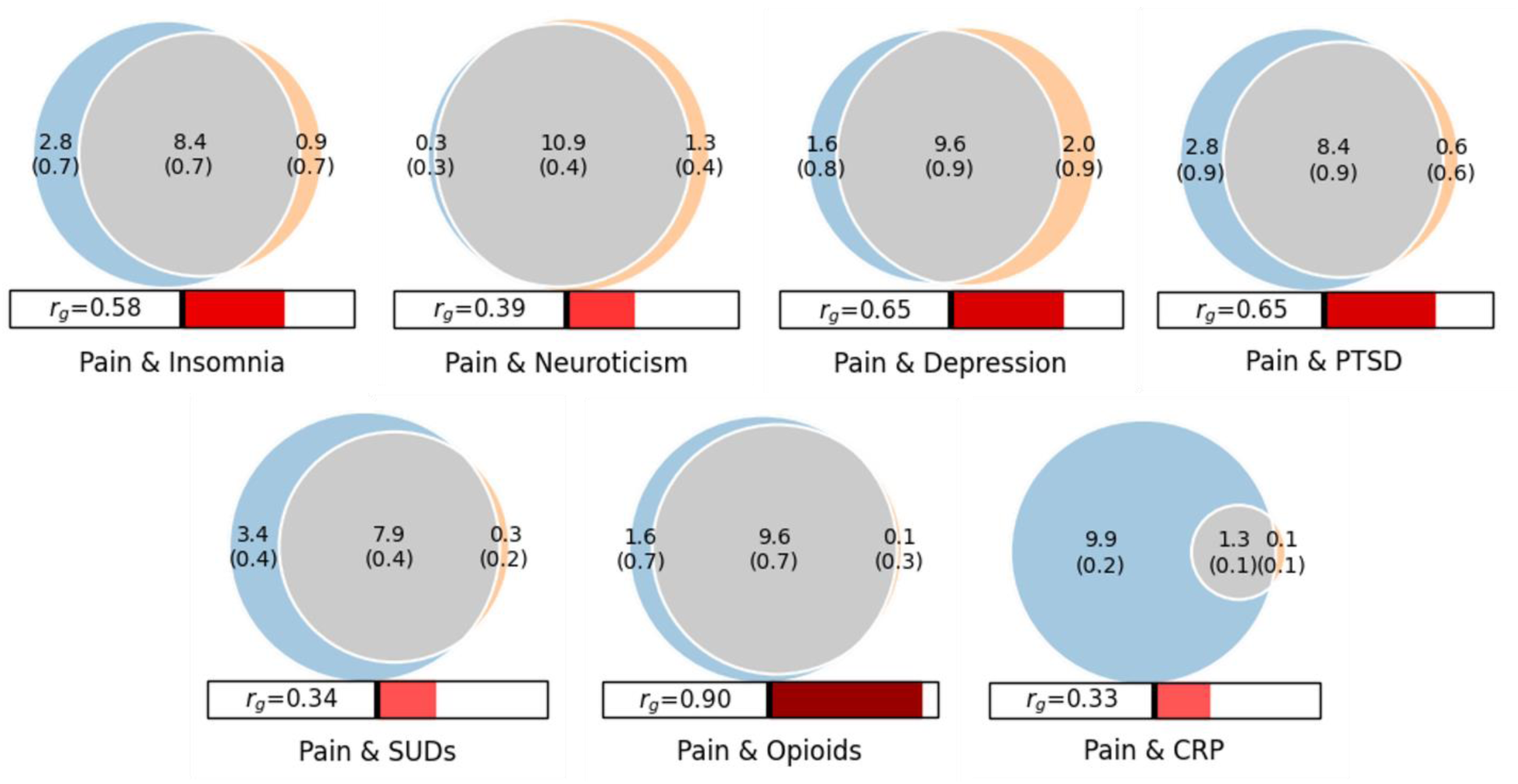
Venn diagrams showing MiXeR results for the estimated number of causal variants for chronic pain shared with psychiatric and substance use disorders, and CRP level. Circles represent shared variants (gray), variants unique to chronic pain (light blue), and variants unique to the other trait of interest (orange). The number of shared variants (and standard deviation) is shown in thousands. Each circle’s size reflects each a trait’s polygenicity, with larger circles corresponding to greater polygenicity. The estimated genetic correlation between chronic pain and each trait from LDSC is shown below the corresponding Venn diagram.

CRP levels had the lowest genetic overlap with chronic pain, with only 20.93% (SD = 0.013) of causal variants shared. However, compared with other traits, CRP levels had far fewer estimated causal variants overall (∼1,400), of which most (∼1,300) were shared with chronic pain. Thus, most of the causal variants for chronic pain (∼9,900) were unique to pain, while the effects of almost all shared variants with CRP level (97.76%) were concordant between the two traits.

### Phenotypic and biomarker pleiotropy for chronic pain

#### PheWAS

Meta-analyzing independent PheWAS data from the Vanderbilt University Medical Center’s biobank (BioVU)^22^ and the Penn Medicine Biobank (PMBB)^23^, we identified associations between chronic pain polygenic scores (PGS) and 202 other phenotypes, including 9 in the neurological domain, 20 brain-related phenotypes, and 174 in other domains (34 circulatory, 14 dermatological, 18 digestive, 14 endocrine/metabolic, 9 genitourinary, 3 hematopoietic, 19 musculoskeletal, 17 respiratory, and 46 other medical conditions) (Extended Data Figure 4, Supplementary Table 22). As expected, the top associations in the neurological domain included acute pain; OR = 1.13, *P* = 1.54 × 10^−20^, back pain; OR = 1.11, *P* = 6.00 × 10^−15^ and other headache syndromes; OR = 1.17, *P* = 4.93 × 10^−16^. Among brain-related phenotypes, notable associations were observed for mood disorders: OR = 1.16, *P* = 9.91 × 10^−61^, depression: OR = 1.15, *P* = 3.28 × 10^−46^, TUD: OR = 1.26, *P* = 4.38 × 10^−21^, PTSD: OR = 1.43, *P* = 3.04 × 10^−20^, and substance addiction and disorders: OR = 1.33, *P* = 6.30 × 10^−26^.

#### LabWAS

After Bonferroni correction, we identified 45 associations of chronic pain PGS with laboratory biomarkers (Extended Data Figure 5, Supplementary Table 23). Consistent with the genetic overlap with CRP levels in the MR and MiXeR analysis, the chronic pain PGS was associated with higher CRP levels (beta = 0.07, SE = 0.008, *P* = 4.72 × 10^−18^). We also identified significant associations with other immune biomarkers (Extended Data Figure 5, Supplementary Table 23), including elevated blood leukocyte (beta = 0.04, SE = 0.004, *P* = 2.04 × 10^−34^) and lymphocyte levels (beta = 0.04, SE = 0.005, *P* = 1.12 × 10^−16^). Other notable associations were with metabolic biomarkers such as higher plasma triglycerides (beta = 0.10, SE = 0.006, *P* = 1.61 × 10^−71^) and lower plasma cholesterol (beta = -0.10, SE = 0.005, *P* = 6.21 × 10^−71^).

## DISCUSSION

In a GWAS meta-analysis of 1,235,695 individuals from three biobanks, we augmented prior findings on the genetic architecture of chronic pain. Specifically, we identified 343 independent loci for chronic pain, including 92 not previously associated with the trait. Using multi-omics gene prioritization analyses, we also highlighted 490 genes associated with chronic pain through brain- and blood-specific regulation. Chronic pain-associated ‘causal’ variants were predicted to contribute substantial proportions of genetic liability for both psychiatric traits and CRP level. Mendelian randomization analyses showed that chronic pain showed potential causal associations with psychiatric disorders, SUDs, CRP level, and cortical area brain structures.

Our results underscore the importance of using large samples to discover new risk loci and biological pathways for chronic pain. For instance, our results revealed strong new signals within chromosome 16 that localized to cytogenetic region p13.3. Here, we found a particularly robust transcriptomic, methylomic, and proteomic association between chronic pain and the *WDR90* gene, which encodes WD repeat-containing protein 90. Converging lines of evidence have linked genetic variation in *WDR90* to insomnia^24^ and substance use^25^ in the general population, with potential mechanisms that include altered expression of WDR90, which can disrupt the structural stability of centrioles^26^. This disruption can lead to aberrant axonal microtubule reorganization and transport, and compromised neural circuitry underlying chronic neuropathic pain^27,28^. We also identified robust transcriptomic and methylomic associations between chronic pain and *DNM1*, which encodes the dynamin-1 protein that plays a key role in recycling synaptic vesicles in the brain, especially the postnatal brain^29^. Variation in this gene has previously been linked to neurodevelopmental disorders of pain sensitivity^30–33^, and changes in the expression or function of dynamin-1 could influence the synaptic regulation in chronic pain as well. Sex differences have been reported in human chronic pain experience^34^ and variations in biological factors could contribute to some of these differences^35,36^. Previous GWAS have identified sex-specific genetic variation for pain^4,5,20,37^. Here, the identification of 115 independent loci, observed in males only, including 12 new pain loci and seven protein-coding genes, highlights the importance of its robust statistical power available by combining MVP and UKB datasets. Notable among the new genetic associations among males are *NXPH2* (chr2q22.1), *SNCA* (chr4q22.1), and *DAAM1* (chr14q23.1). *NXPH2* encodes neurexophilin 2, a member of the neurexophilin family, and modulates synaptic plasticity in interneurons critical for pain perception^38,39^. *SNCA* encodes α-synuclein, a small (140 amino acid) protein localized, in part, to presynaptic terminals that modulate vesicle trafficking and neurotransmitter release^40^. *DAAM1* encodes disheveled associated activator of morphogenesis 1, involved in presynaptic actin assembly^41^, with upregulated miRNA expression of the gene linked with chronic painful neuropathy^42^. We also identified 12 independent loci among females, including two – *MAML3* (q31.1) and *GABRB2* (q34) – not previously identified in the contributing GWASs or among males. *MAML3* encodes mastermind-like transcriptional coactivator 3, involved in the notch signaling pathway, which aids neural development and function^43^. *GABRB2* encodes the beta-2 subunit of the GABA-A receptor, which is involved in regulating neuronal excitability and maintaining the balance between neuronal excitation and inhibition^44^.

There is overlap of physical, emotional, and behavioral supraspinal pain pathways^45^, with both biological and psychosocial factors contributing to the etiology of chronic pain. Consistent with this, chronic pain is highly comorbid with psychiatric disorders, such that individuals with psychiatric disorders experience more severe pain, with poorer functioning, greater disability, and a higher incidence of opioid addiction^46^. The most prevalent psychiatric comorbidities associated with chronic pain are anxiety, depression, and insomnia^47^, with observational studies suggesting a bidirectional causal relationship of chronic pain with these conditions^48–51^. Consistent with findings from previous MR studies of chronic widespread pain^52^ and pain localized at different bodily sites^53,54^, we found a bidirectional association between chronic pain and MDD, PTSD, and PTSD symptom clusters. We previously reported causal relations between pain intensity and SUDs^5^, in line with others^55,56^ and consistent with the finding here of a causal effect of chronic pain on SUDs (alcohol, cannabis, opioids, and tobacco) and shared causal variants with opioid use and a SUD latent factor. In line with prior reports^54^, chronic pain was also associated with an elevated risk of insomnia. The identification of shared neural circuits that provide mechanistic explanations for these causal associations could be important clinically insofar as interventions that target these circuits could have beneficial effects both on the experience of pain and psychiatric symptoms.

Changes in brain structure and function that accompany the chronification of pain^45^, informed by neuroimaging studies^16^, can help to localize brain regions associated with chronic pain. Genetic variants pleiotropic for multisite chronic pain and SUDs are associated with multiple cerebellar and amygdalar imaging phenotypes^55^ and suggest a role for subcortical shared pain-addiction pathogenetic pathways. It has been reported that lower cortical surface area activity plays a role in chronic pain pathology^12^, while others have observed that loci affecting regional cortical surface area clusters are associated with psychiatric disorders^57,58^. By integrating our GWAS results with brain structure IDPs, we extend these findings to include chronic pain, where we show that allelic variation associated with a smaller cortical surface area and greater cortical thickness is also present in chronic pain. Using MR, we observed a causal effect of lower cortical surface area, particularly in the right and left hemispheres, on increased risk for chronic pain. Together with evidence of a shared genetic basis for reduced gray matter of the insula and pain at different bodily sites^59^, these findings corroborate observational reports of changes in brain gray matter in the setting of chronic pain^12,60^. These IDPs may serve as preclinical indicators of the risk of chronic pain and, if the findings are independently validated using neuroimaging data^16^, could help in the prevention or early diagnosis of chronic pain among individuals who experience clinically significant acute pain.

Inflammatory mechanisms may play a role in the development and maintenance of pain^61^. CRP, an immune regulator with potential causal roles in inflammatory musculoskeletal conditions^62,63^, is one of the most studied biomarkers in non-inflammatory pain. However, MR studies of the association of CRP with such conditions have yielded mixed results^61,64–66^. A recent study showed no causal link between CRP and multisite chronic pain (*n* = 387,649), chronic widespread pain (*n* = 249,843), or spinal pain (*n* = 1,028,947)^66^, possibly due to the use of a case-control study design or the pain phenotypes included. Here, we found several lines of evidence supporting the link between CRP and chronic pain, including a causal effect of chronic pain on CRP level, evidence of overlapping causal genetic variants for chronic pain and CRP level, and an association between chronic pain PGS and elevated CRP level in a LabWAS. These findings are consistent with previous reports of higher CRP levels in chronic back pain^64^ and fibromyalgia^63^, though the potential utility of targeting immune-related biomarkers for pain as a therapeutic strategy remains to be demonstrated.

Study limitations include an exclusively EUR-ancestry sample, which limits the application of the study’s findings to non-EUR populations. To permit well-powered meta-analyses of chronic pain that adequately represent genetically diverse populations, greater availability of large GWAS of pain traits is needed in non-EUR samples. While the genetic correlation among pain phenotypes included in the meta-analysis was significant and positive, they were far from unity, likely due to the heterogeneity of the pain phenotypes resulting from different methods of assessing pain across the biobanks (i.e., quantitatively in MVP and UKB, and as a binary measure in FinnGen). Whereas MVP and UKB pain phenotypes are ordinal measures that reflect the chronicity of pain, the FinnGen pain phenotype is a binary measure of general pain across different body regions that does not specify a timeframe. By conducting a secondary GWAS in which the FinnGen sample was excluded, we confirmed a preponderance (∼73%) of pain-associated loci and observed similar genetic correlation patterns to complex traits as in the primary analyses. The secondary analyses also identified 14 new pain associations.

Chronic pain is a highly variable condition due to variations in pain tolerance, the use of different pain assessments, and the effects of chronic pain treatments^67^. Refined phenotyping that explores similarities and differences across pain conditions is needed to elucidate common treatment targets^16^. Therefore, future GWASs should prioritize deep phenotyping initiatives that facilitate a more systematic evaluation of the sources of pain and consistencies across pain assessments in multi-biobank samples.

Despite these limitations, the substantially larger sample size and statistical power of this meta-analysis identified many new loci associated with the risk of chronic pain. Our multi-omics analytic approach highlighted effector genes that could serve as targets for new therapeutics. The converging lines of evidence across multiple analytic methods that elucidate the pleiotropic associations of pain-related loci with psychiatric, SUD, immune traits, and neural structures enhance the validity of the findings. This approach highlights the potential utility of the GWAS data for evaluating the impact of genetic risk for chronic pain on multiple traits of clinical interest and paves the way for biomarker development strategies that could be applied to chronic pain and the many traits commonly associated with it.

## METHODS

### GWAS data selection

We included summary statistics from prior published GWAS conducted in three large biobanks (Supplementary Table 1)^3,5,18^. The first GWAS was performed by us in the Million Veteran Program (MVP) (*N* = 436,683 individuals of European-like ancestry (EUR))^5^. This study examined pain intensity (as a proxy for chronic pain), represented by the median of the annual median pain ratings measured across multiple years with an 11-point ordinal numeric rating scale, a consistent and valid measure of self-reported pain^68,69^. The second GWAS was conducted in the UKB cohort (*N* = 387,649 EUR individuals)^3^. That study examined multisite chronic pain (MCP), defined as the sum of chronic pain across 7 bodily sites (i.e., head, face, neck/shoulder, back, stomach/abdomen, hip, knee), which yields an 8-point ordinal score. Finally, we used data from a GWAS of “pain (limb, back, neck, head abdominally)” ascertained using ICD-9 and ICD-10 codes (Finnish version) from the FinnGen cohort (data freeze 10; *N* = 411,363 EUR individuals)^18^. The FinnGen study assessed overall pain, with cases (*N* = 189,683) required to have one or more ICD-9 or -10 diagnostic codes for 16 disorders with a pain component (occurring in joints, limbs, neck, head, abdomen, and back) and none for controls (*N* = 221,680) (see details in Supplementary Table 1).

Because these three samples are largely comprised of EUR individuals, we limited the analyses to that population group. The primary meta-analysis used data from all three GWAS samples (*N* = 1,235,695). We also conducted a secondary meta-analysis in a subsample of 851,332 individuals that excluded participants from FinnGen to assess potential confounding by differences in pain assessments and the founder effect in that cohort^18^. Furthermore, because sex-specific information is available only for the MVP pain intensity (male; *N* = 404,510 and female; *N* = 32,173)^5^ and UKB MCP (male; *N* = 178,556 and female; *N* = 209,093)^20^ GWAS, our sex-specific analyses were limited to those two samples.

### Association analyses and risk locus definition

We calculated genetic correlations among the three pain phenotypes using LDSC and conducted meta-analyses using a sample-size weighted method in METAL^70^. For uniformity, we included only variants with a minor allele frequency >1% in both primary and sex-stratified meta-analyses. Variants with *P* < 5 × 10^−8^ were considered GWS. With LDSC attenuation ratios close to 0 and none of the GWS lead single-nucleotide polymorphisms (SNPs) showing evidence of heterogeneity across cohorts, we did not use the genomic control option in METAL.

To identify risk loci and their lead variants, we performed LD clumping in FUMA^71^ at a range of 3,000 kb, *r*^2^ > 0.1, using the EUR1000 Genomes ancestry reference panel^72^. Following clumping, genomic risk loci within +/- 1 MB of one another were incorporated into the same locus. We used GCTA-COJO^73^ to define independent variants by conditioning them on the most significant variant within the locus, with significant variants (*P* < 5 × 10^−8^) after conditioning considered to be independently associated with the phenotype.

We determined which of the independent variants were credible by merging risk variants within +/- 1 Mb of the lead variant and fine-mapping the resulting region with 95% credible sets using FINEMAP^74^. Posterior inclusion probabilities (PP) range from 0 to 1, with values closer to 1 indicating greater causal probability for the variant. We implicated a putative causal variant if it accounted for >50% of the PP in the 95% credible set.

### Prioritization of effector genes for chronic pain

We used MAGMA^75^ in FUMA (v1.3.6a)^71^ to map chronic pain GWAS SNPs to 19,437 protein-coding genes according to their physical position in NCBI build 37 and performed gene ontology analyses to identify biological processes and pathways associated with the trait.

To prioritize potential causal genes at identified chronic pain risk loci, we integrated the GWAS results with multi-omics data using Summary-based Mendelian Randomization (SMR)^76^ and chromatin interaction (Hi-C) coupled MAGMA (H-MAGMA)^77^. In SMR analysis, we examined effects on gene expression using expression quantitative trait locus (eQTL) data from BrainMeta^78^ and GTEx v.8^79^; quantified splicing using sQTL data from BrainMeta^78^; brain and blood methylation levels using mQTL data from BrainMeta^78^ (brain mQTL) and McRae et al.^80^ (blood mQTL), respectively; plasma protein abundance levels using pQTL data from deCODE^81^ and UKB^82^; and brain protein abundance levels using pQTL data from Wingo et al^83^. We used H-MAGMA^77^ to assign noncoding (intergenic and intronic) SNPs to genes based on their chromatin interactions. H-MAGMA uses six Hi-C datasets from human cortical tissue across two developmental stages (prenatal and postnatal) and brain cell types (iPSC-derived neurons and astrocytes), enabling development- and cell-type-specific gene mapping. We applied the HEIDI test^84^ to remove SMR signals (*P*_HEIDI_ < 0.05) that were due to LD between pain-associated variants and eQTLs/sQTLs/mQTLs/pQTLs. We applied a Bonferroni correction for all genes tested in each omics analysis (*α* = 0.05/*n* genes tested).

To generate a ranked list of effector genes for use as candidate drug targets for chronic pain, we combined evidence across the omics analyses (i.e., SMR and H-MAGMA). We binarized (yes=1, no=0) and then summed the evidence supporting a particular gene. Each gene was then ranked based on its summed prioritization score. For example, if chronic pain was significantly associated with a gene locus across eQTL (+1), sQTL (+1), Hi-C chromatin interaction (+1), mQTL (+1), blood pQTL (+1), and brain pQTLs (+1) analyses, the summed score for the locus would be 6. Note, that this approach is biased toward proteins (or protein-coding genes), as they have a greater scoring potential.

### Factor structure of pain intensity and other chronic pain conditions

We used genomic structural equation modeling (gSEM)^85^ to evaluate how pain intensity aligns with an established factor model of other chronic pain conditions^7^. First, we examined genetic correlations between pain intensity and 33 heritable chronic pain conditions from the UKB using LDSC implemented in the GenomicSEM package. Next, replicating the previously identified factor structure across chronic pain conditions^7^, we fit two nested CFAs: one incorporating pain intensity within both the general pain factor and the musculoskeletal pain factor, and one with pain intensity loading solely on the general pain factor. We used a chi-square difference test, which leverages the models’ nested structure, to determine whether adding pain intensity to the musculoskeletal pain factor significantly improved model fit. Model fit was also evaluated by comparing the AIC, where lower is better, CFI; >0.9, and SRMR; <0.08 values.

### Genetic correlation with complex traits and brain structure

We used LDSC^19^ and publicly available GWAS data to perform batch genetic correlations between chronic pain, biological variables and complex disease traits (*n* = 80), and brain IDPs with significant heritability (*n* = 1,323)^21^. The 1000 Genomes Project European populations served as the LD reference panel (version 3),^72^ consistent with the ancestry of the meta-GWAS sample. We applied a Bonferroni correction (*α* = 0.05/*n* traits tested) to account for multiple testing and identify significant correlations.

### Genetically informed causal analysis

We performed bi-directional, two-sample MR analyses using the TwoSampleMR R package (v0.5.6)^86^ to test causal relations between traits genetically correlated with chronic pain. We defined genetic instruments using independent GWS variants (*P* < 5 × 10^−8^; LD clumping criteria: r^2^ = 0.001 and a 10,000-kb window). For traits with <10 GWS variants after clumping, we used a suggestive threshold of *P* < 5 × 10^−6^ to select instrumental variables (IVs) for MR.

We included only variants present in both the exposure and outcome datasets. To quantify the strength of IVs, we calculated the *F*-statistics of all genetic instruments using the per-allele effect size of the SNP association with the phenotype (*β*) and s.e. using the formula^87^ *F*-statistic = (*β*/s.e.)^2^. IVs with *F*-statistic estimates <10 were considered weak instruments that could bias results^88^. We used Steiger’s test^89^ to determine whether the SNP–outcome correlation is greater than the SNP–exposure correlation. Because SNPs that fail Steiger’s test may not be primarily associated with the exposure (Steiger, *P* > 0.05), they were filtered out before the MR analysis. Whereas horizontal pleiotropy can bias MR findings, we assessed its presence by measuring the SNPs’ heterogeneity using the *I*^2^ index and Cochran’s *Q* heterogeneity test. When significant heterogeneity was observed, we used the MR–Egger intercepts to assess the IV estimate’s reliability and test for horizontal pleiotropy.

MR estimates were generated using inverse variance weighted (IVW), MR-Egger, and weighted median^86^ tests. For complex traits, potential causal effects were those for which at least two MR tests were significant after multiple correction (Bonferroni *P* = 0.05/ *n* traits tested). Because brain IDP pairs measured in the same brain region are highly correlated^90^, causal associations with brain IDPs were determined using one MR test to avoid overly conservative correction. We tested for the absence of horizontal pleiotropy using MR–Egger intercept (*P* > 0.05).

### Characterizing polygenic pleiotropy

We used MiXeR software to conduct univariate and bivariate causal mixture models^91,92^. Univariate causal mixture models use GWAS summary statistics to estimate a trait’s polygenicity (i.e., the number of causally associated variants needed to explain 90% of the SNP-heritability) and discoverability (i.e., the effect size variance). Univariate MiXeR models that yield positive AIC values are sufficiently powered for bivariate causal mixture models.

In contrast to genetic correlations, bivariate MiXeR models estimate the genetic overlap between two traits, irrespective of the direction of the effect of the causal variant on the trait. After ensuring that there was sufficient power, we used bivariate MiXeR models to estimate the degree of genetic overlap between chronic pain and CRP levels^93^, depression^94^, insomnia^24^, neuroticism^95^, opioid medication use^96^, PTSD^97^, and a SUD latent factor^98^. These traits were selected for their biological and clinical relevance to chronic pain, including their potential to share genetic pathways with it. We used Dice coefficients to evaluate the proportion of causal variants shared by each pair of traits.

### Phenome- and lab-wide associations for chronic pain

We conducted PheWAS using chronic pain PGS in two independent datasets: Vanderbilt University Medical Center’s biobank (BioVU)^22^ and the University of Pennsylvania’s Penn Medicine Biobank (PMBB)^23^.

*BioVU:* The BioVU cohort comprises Vanderbilt University Medical Center patients with electronic health record (EHR) and genotype data, all with informed consent^99^. Genotyping was performed using the Illumina Multi-Ethnic Genotype Array (MEGAEX), as previously described^22^. Genotypes were filtered for SNP (< 0.95) and individual (< 0.98) call rates, sex discrepancies, and excessive heterozygosity (|Fhet| > 0.2)^100^. Principal component analysis (PCA) was performed using FlashPCA2 1000 Genomes phase 3 reference datasets^72^ to identify EUR individuals. Genotypes were imputed using the Michigan Imputation Server (https://imputationserver.sph.umich.edu) and the Haplotype Reference Consortium panel^101^. SNPs with imputation quality r^2^ > 0.3 or INFO > 0.95 and MAF > 0.01 were retained for PGS analyses.

*PMBB*: The PMBB cohort comprises Penn Medicine patients with EHR phenotypes and genotype data, with informed consent obtained from all participants^23^. Genotyping was performed using the GSA genotyping array. Genotype phasing was performed using EAGLE^102^. PCA in the smartpca module of the Eigensoft package (https://github.com/DReichLab/EIG) was used to determine the genetic ancestry of EUR subsets by mapping the data onto the 1000 Genomes reference panel^72^. Genotypes were imputed to the TOPMed imputation reference panel^103^ and imputation dosages were converted to best-guess hard-called genotypes. We retained SNPs with INFO > 0.7, genotype call rate > 0.99, sample call rate > 0.99, and MAF > 0.01 for PGS analyses.

We calculated PGS for the chronic pain GWAS summary data in BioVU and PMBB using PRS-CS software,^104^ applying the default settings to estimate shrinkage parameters. ICD-9 and ICD-10 codes from EHR data or diagnostic interviews were mapped to phecodes. PheWAS were conducted using logistic regression models in the PheWAS v0.12 R package^105^, with age, sex, and the first 10 genetic ancestry principal components as covariates. PheWAS were performed on 66,917 and 29,355 individuals in the BioVU and PMBB cohorts, respectively. PheWAS results for 1,817 unique phenotypes across both datasets were meta-analyzed by calculating a weighted average of each cohort’s beta estimates and standard errors.

We also performed LabWAS in BioVU to examine the association between chronic pain PGS and 315 lab test results and/or biomarkers^22^. For both PheWAS and LabWAS, Bonferroni correction (0.05/*n* traits) was applied to identify significant associations.

## Data Availability

The full summary statistics from the meta-analyses will be available upon request. Each of the summary statistics included in the meta-analyses are publicly available.^3,5,18,20^

## Code Availability

Meta-analyses were performed using METAL (https://genome.sph.umich.edu/wiki/METAL_Documentation). GCTA (https://cnsgenomics.com/software/gcta/#Overview) was used for the identification of independent loci (GCTA-COJO). FINEMAP (http://www.christianbenner.com/) was used to fine-map genomic risk loci. FUMA (https://fuma.ctglab.nl/) was used for gene-set enrichment analyses. Gene prioritization was performed using SMR (https://yanglab.westlake.edu.cn/software/smr/#Overview) and H-MAGMA (https://github.com/thewonlab/H-MAGMA). LDSC (https://github.com/bulik/ldsc) was used for heritability estimation and genetic correlation analysis. Polygenic overlap was determined using MiXeR (https://github.com/precimed/mixer). PRS analyses were performed using PRS-CS (https://github.com/getian107/PRScs). PheWAS analyses were run using the PheWAS R package (https://github.com/PheWAS/PheWAS). The MendelianRandomization R package (https://cran.r-project.org/web/packages/MendelianRandomization/index.html) was used for MR analyses. Genomic SEM was conducted using the GenomicsSEM R package (https://github.com/GenomicSEM/GenomicSEM).

## Acknowledgements

This work was supported by Merit Review Awards from the US Department of Veterans Affairs Biomedical Laboratory Research and Development Service (no. I01 BX003341 (to H.R.K.)) and Clinical Science Research and Development Service (no. I01 CX001734 (to K.M.K.)); NIH R01 MH113362 (to L.K.D); Tobacco-Related Disease Research Program grant 32IR5226 (to S.S-R); the VISN 4 Mental Illness Research, Education and Clinical Center (to H.R.K.); NIAAA grant K01 AA028292 (to R.L.K.); and NIDA grant DA046345 (to H.R.K.). The funders had no role in study design, data collection and analysis, decision to publish, or preparation of the manuscript.

We acknowledge the Penn Medicine BioBank (PMBB) for providing data for generating polygenic risk scores and PheWAS analyses and thank the patient-participants of Penn Medicine who consented to participate in this research program. We would also like to thank the Penn Medicine BioBank team and Regeneron Genetics Center for providing genetic variant data for analysis. The PMBB is approved under IRB protocol# 813913 and supported by Perelman School of Medicine at the University of Pennsylvania, a gift from the Smilow family, and the National Center for Advancing Translational Sciences of the National Institutes of Health under CTSA award number UL1TR001878.

## Contributions

S.T. conducted the main analyses and drafted the manuscript. C.D., Z.J., Y.K., and M.J. provided analyses. C.D. helped in writing the manuscript. R.L.K. supervised the analyses and helped in writing the manuscript. H.R.K. conceived the project, obtained funding to support it, and helped supervise the analyses and write the manuscript. All authors reviewed and approved the final version of the manuscript.

## Ethics declarations

Dr. Kranzler is a member of advisory boards for Altimmune and Clearmind Medicine; a consultant to Sobrera Pharmaceuticals and Altimmune; the recipient of research funding and medication supplies for an investigator-initiated study from Alkermes; a member of the American Society of Clinical Psychopharmacology’s Alcohol Clinical Trials Initiative, which was supported in the last three years by Alkermes, Dicerna, Ethypharm, Imbrium, Indivior, Kinnov, Lilly, Otsuka, and Pear; and an inventor on U.S. provisional patent “Multi-ancestry Genome-wide Association Meta-analysis of Buprenorphine Treatment Response.”

---

**Extended Data Figure 1.**
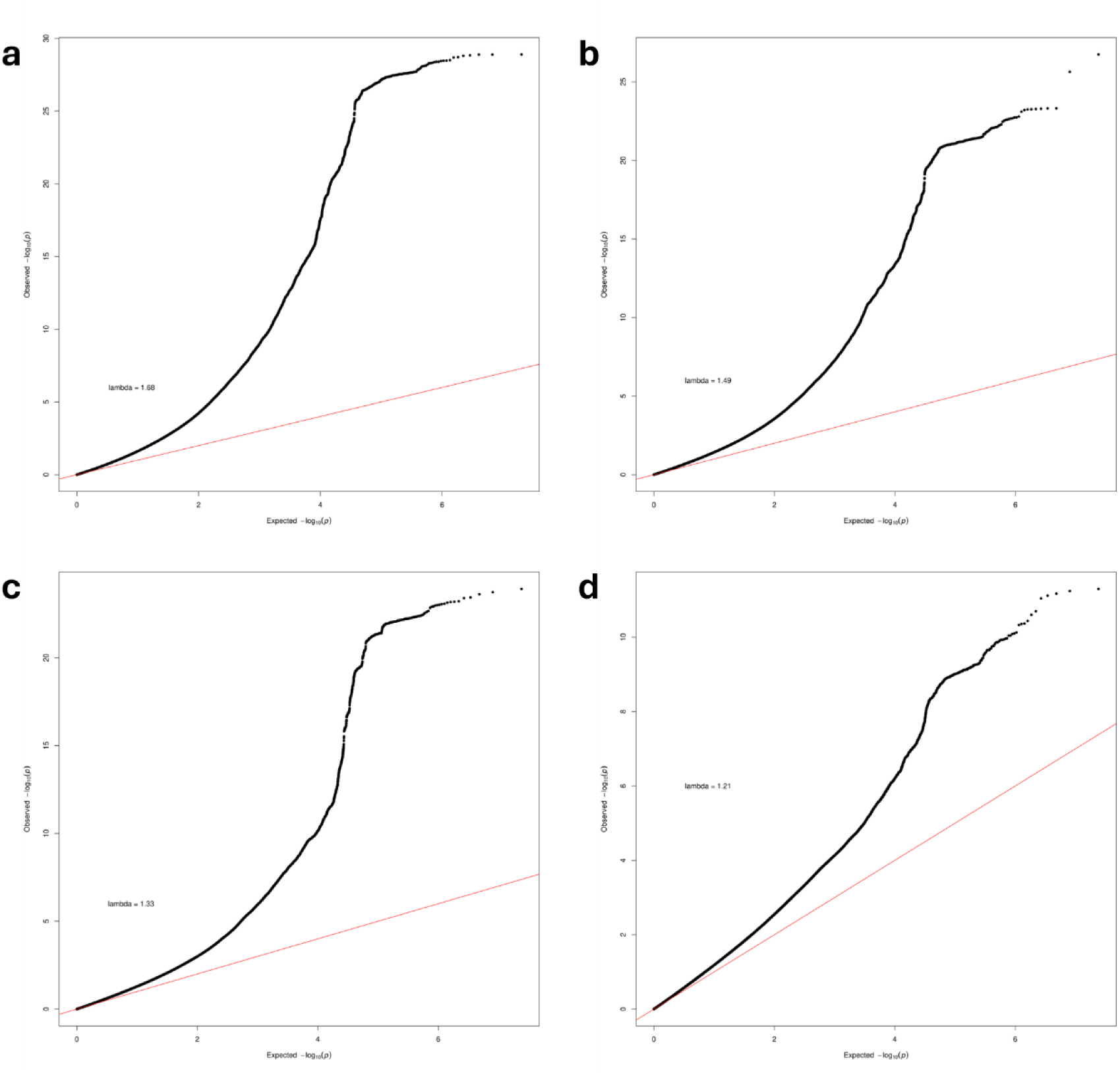
Quantile–quantile (QQ) plots of the SNP-based associations with chronic pain in the (a) primary (b) secondary (c) male and (c) female GWASs. QQ plots show inflated lambda values (primary; λ_GC_ = 1.68, secondary; λ_GC_ = 1.49, male; λ_GC_ = 1.33, and female; λ_GC_ = 1.21), however, the LDSC intercept (1.1; standard error (s.e.) = 0.01) and attenuation ratio (0.03 – 0.10) (Supplementary Table 3) both indicate that the inflation is largely due to true polygenicity. SNP P values were computed in METAL using a two-sided, sample-size-weighted Z-score method. QQ plot estimates were generated using a chi-squared test.

**Extended Data Figure 2.**
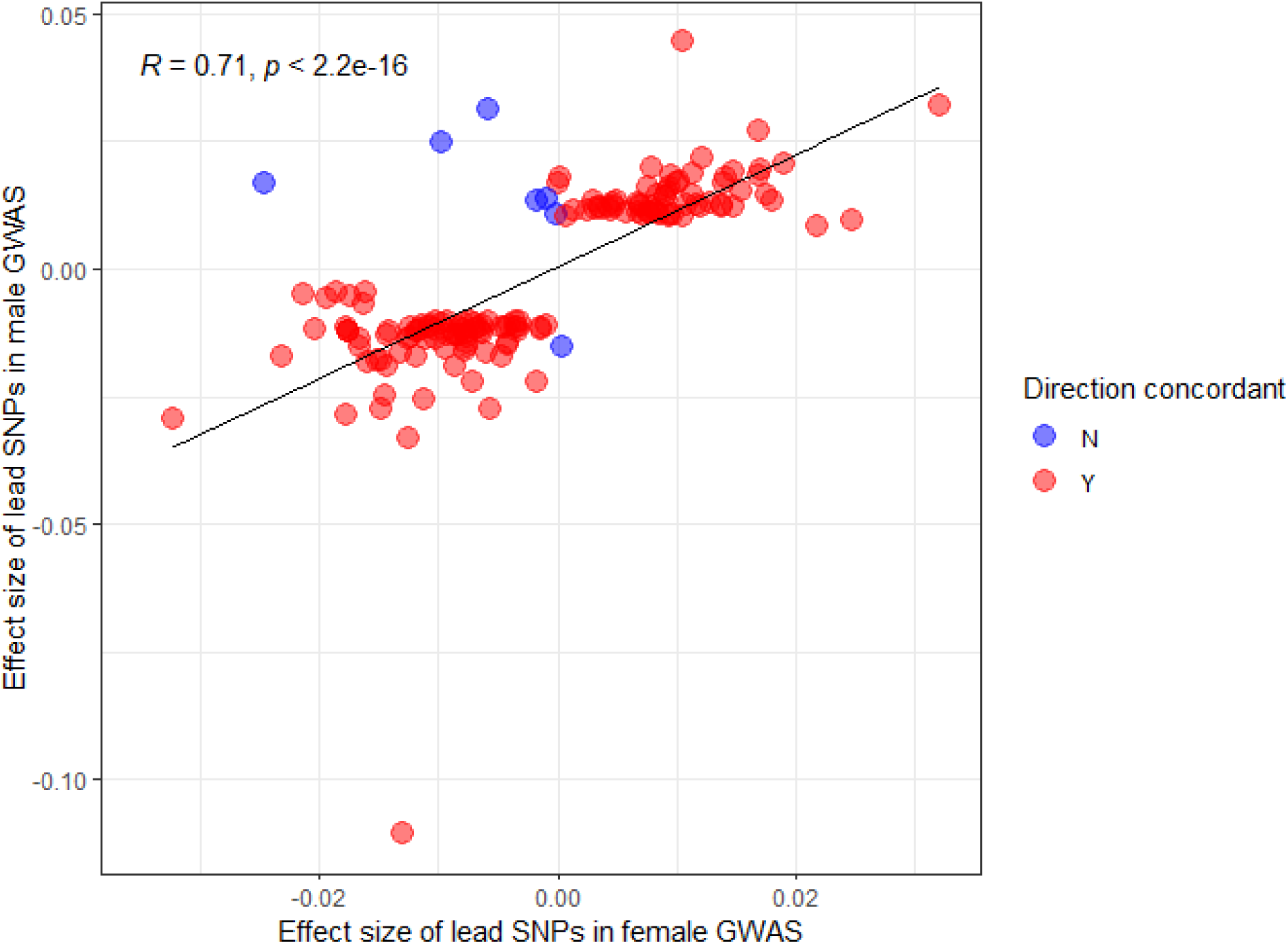
The magnitude and direction of the effect sizes are plotted for each GWAS. The results show a significant (*P* < 2.2 × 10^−16^) high correlation between the effect sizes (β) of chronic pain lead SNPs for male and female meta-GWASs.

**Extended Data Figure 3.**
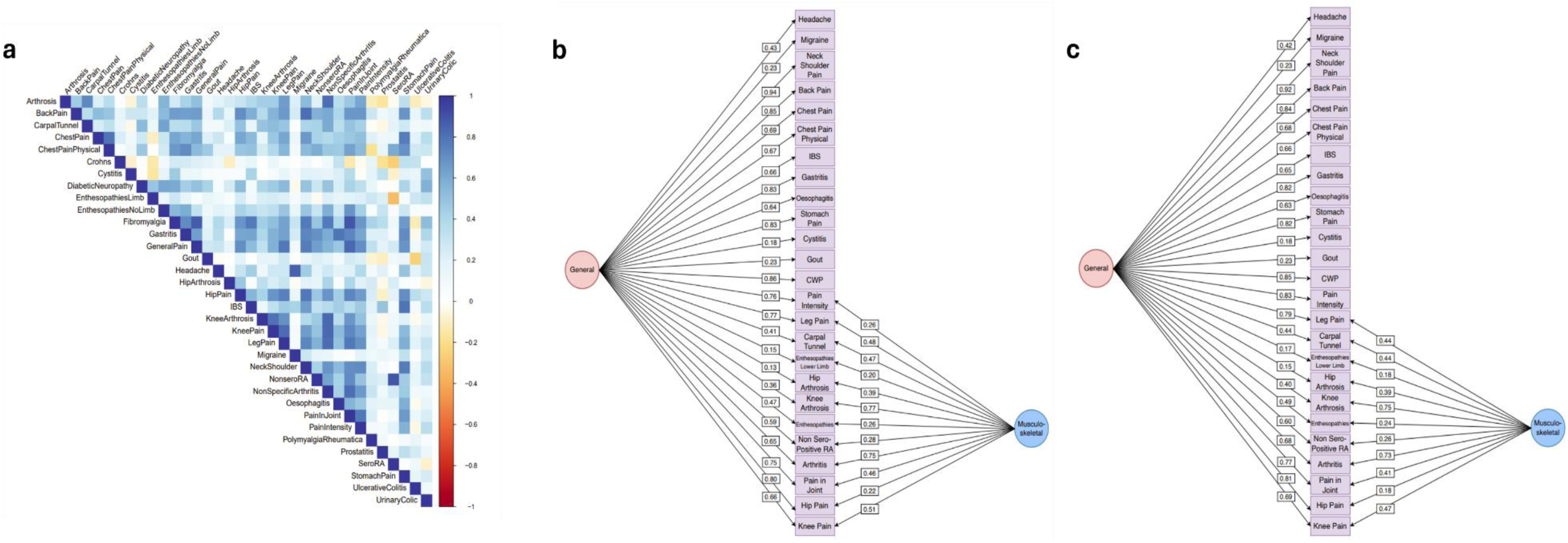
Genomic structural equation modeling showing the genetic relationship of pain intensity and other pain conditions in the UKB. (a) Heatmap showing genetic correlation across pain conditions. Chronic pain factor showing EFA-CFA model for 26 pain conditions with SNP effects where pain intensity loads to (b) both a general and musculoskeletal factor and (c) only the general pain factor.

**Extended Data Figure 4:**
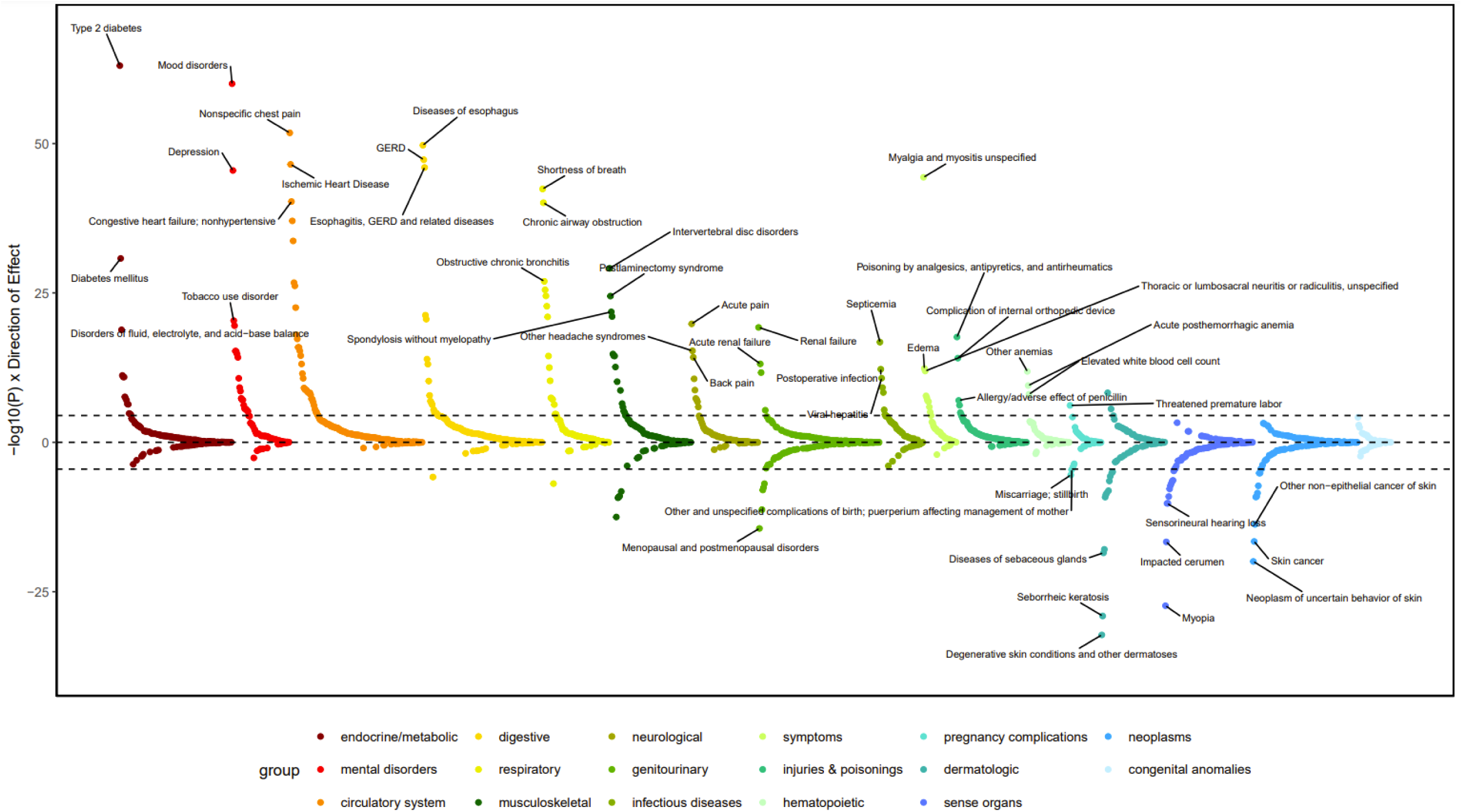
PheWAS meta-analysis results showing phenotypic associations. PheWAS plot for chronic pain PGS in EUR individuals from PMBB and BioVU. The top four phenotypes that pass multiple testing correction in each phenotypic grouping (black dashed line) are annotated (Bonferroni correction threshold = *P* < 2.75 × 10^-5^ (0.05/1,817). See Supplementary Table 21 for full Bonferroni-significant results.

**Extended Data Figure 5.**
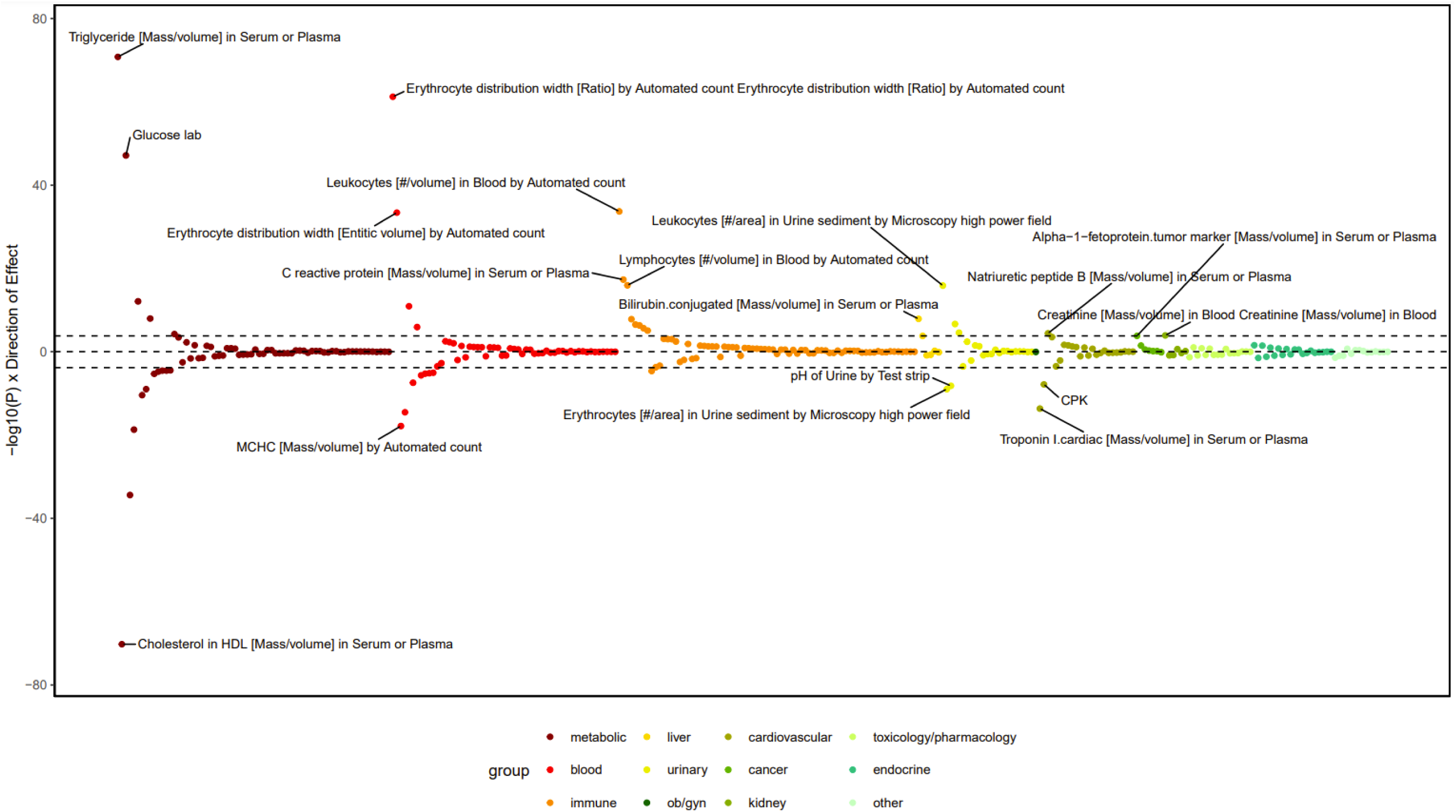
LabWAS results showing laboratory measure associations. LabWAS plot for chronic pain PGS in EUR individuals from BioVU. The top three phenotypes that pass multiple testing correction in each laboratory grouping (black dashed line) are annotated (Bonferroni correction threshold = *P* < 1.59 × 10^-4^ (0.05/315). See Supplementary Table 22 for full Bonferroni-significant results.

## Notes

### Funding Statement

This study was supported by Merit Review Awards from the US Department of Veterans Affairs Biomedical Laboratory Research and Development Service (no. I01 BX003341 (to H.R.K.)) and Clinical Science Research and Development Service (no. I01 CX001734 (to K.M.K.)); NIH R01 MH113362 (to L.K.D); Tobacco-Related Disease Research Program grant 32IR5226 (to S.S-R); the VISN 4 Mental Illness Research, Education and Clinical Center (to H.R.K.); NIAAA grant K01 AA028292 (to R.L.K.); and NIDA grant DA046345 (to H.R.K.). The funders had no role in study design, data collection and analysis, decision to publish, or preparation of the manuscript.
We acknowledge the Penn Medicine BioBank (PMBB) for providing data for generating polygenic risk scores and PheWAS analyses and thank the patient-participants of Penn Medicine who consented to participate in this research program. We would also like to thank the Penn Medicine BioBank team and Regeneron Genetics Center for providing genetic variant data for analysis. The PMBB is approved under IRB protocol# 813913 and supported by Perelman School of Medicine at the University of Pennsylvania, a gift from the Smilow family, and the National Center for Advancing Translational Sciences of the National Institutes of Health under CTSA award number UL1TR001878.

### Author Declarations

Ethics committee of the University of Pennsylvania Penn Medicine Biobank and Vanderbilt University Medical Center biobank gave ethical approval for this work.

## REFERENCES

1. Cohen, S. P., Vase, L. & Hooten, W. M. Chronic pain: an update on burden, best practices, and new advances. The Lancet 397, 2082–2097 (2021).

2. Ray, B. M., Kelleran, K. J., Fodero, J. G. & Harvell-Bowman, L. A. Examining the relationship between chronic pain and mortality in U.S. Adults. J. Pain 25, (2024).

3. Johnston, K. J. A. et al. Genome-wide association study of multisite chronic pain in UK Biobank. PLOS Genet. 15, e1008164 (2019).

4. Mocci, E. et al. Genome wide association joint analysis reveals 99 risk loci for pain susceptibility and pleiotropic relationships with psychiatric, metabolic, and immunological traits. PLoS Genet. 19, e1010977 (2023).

5. Toikumo, S. et al. A multi-ancestry genetic study of pain intensity in 598,339 veterans. Nat. Med. 30, 1075–1084 (2024).

6. Maixner, W., Fillingim, R. B., Williams, D. A., Smith, S. B. & Slade, G. D. Overlapping Chronic pain conditions: Implications for diagnosis and classification. J. Pain 17, T93– T107 (2016).

7. Zorina-Lichtenwalter, K. et al. Genetic risk shared across 24 chronic pain conditions: identification and characterization with genomic structural equation modeling. PAIN 164, (2023).

8. Tanguay-Sabourin, C. et al. A prognostic risk score for development and spread of chronic pain. Nat. Med. 29, 1821–1831 (2023).

9. Apkarian, A. V., Baliki, M. N. & Geha, P. Y. Towards a theory of chronic pain. Prog. Neurobiol. 87, 81–97 (2009).

10. Barroso, J., Branco, P. & Apkarian, A. V. Brain mechanisms of chronic pain: critical role of translational approach. Transl. Res. 238, 76–89 (2021).

11. Kuner, R. & Flor, H. Structural plasticity and reorganisation in chronic pain. Nat. Rev. Neurosci. 18, 20–30 (2017).

12. Bhatt, R. R. et al. Mapping brain structure variability in chronic pain: the role of widespreadness and pain type and its mediating relationship with suicide attempt. Biol. Psychiatry 95, 473–481 (2024).

13. Davis, K. D. et al. Discovery and validation of biomarkers to aid the development of safe and effective pain therapeutics: challenges and opportunities. Nat. Rev. Neurol. 16, 381–400 (2020).

14. Eldabe, S., Obara, I., Panwar, C. & Caraway, D. Biomarkers for chronic pain: Significance and summary of recent advances. Pain Res. Manag. 2022, 1940906 (2022).

15. Fillingim, M. et al. A Biomarker-centric framework for the prediction of future chronic pain. medRxiv 2024.04.19.24306101 (2024) doi:10.1101/2024.04.19.24306101.

16. Quidé, Y. et al. ENIGMA-Chronic Pain: a worldwide initiative to identify brain correlates of chronic pain. PAIN 165, (2024).

17. Farrell, S. F. et al. A shared genetic signature for common chronic pain conditions and its Impact on Biopsychosocial Traits. J. Pain 24, 369–386 (2023).

18. Kurki, M. I. et al. FinnGen provides genetic insights from a well-phenotyped isolated population. Nature 613, 508–518 (2023).

19. Bulik-Sullivan, B. K. et al. LD Score regression distinguishes confounding from polygenicity in genome-wide association studies. Nat. Genet. 47, 291–295 (2015).

20. Johnston, K. J. A. et al. Sex-stratified genome-wide association study of multisite chronic pain in UK Biobank. PLoS Genet. 17, e1009428 (2021).

21. Smith, S. M. et al. An expanded set of genome-wide association studies of brain imaging phenotypes in UK Biobank. Nat. Neurosci. 24, 737–745 (2021).

22. Dennis, J. K. et al. Clinical laboratory test-wide association scan of polygenic scores identifies biomarkers of complex disease. Genome Med. 13, 6 (2021).

23. Verma, A. et al. The Penn Medicine BioBank: Towards a genomics-enabled learning healthcare system to accelerate precision medicine in a diverse population. J. Pers. Med. 12, (2022).

24. Jansen, P. R. et al. Genome-wide analysis of insomnia in 1,331,010 individuals identifies new risk loci and functional pathways. Nat. Genet. 51, 394–403 (2019).

25. Xu, K. et al. Genome-wide association study of smoking trajectory and meta-analysis of smoking status in 842,000 individuals. Nat. Commun. 11, 5302 (2020).

26. Steib, E. et al. WDR90 is a centriolar microtubule wall protein important for centriole architecture integrity. eLife 9, e57205 (2020).

27. Lindhout, F. W. et al. Centrosome-mediated microtubule remodeling during axon formation in human iPSC-derived neurons. EMBO J. 40, e106798 (2021).

28. Finnerup, N. B., Kuner, R. & Jensen, T. S. Neuropathic pain: from mechanisms to treatment. Physiol. Rev. 101, 259–301 (2021).

29. Ferguson, S. M. & De Camilli, P. Dynamin, a membrane-remodelling GTPase. Nat. Rev. Mol. Cell Biol. 13, 75–88 (2012).

30. von Spiczak, S. et al. DNM1 encephalopathy. Neurology 89, 385–394 (2017).

31. Jones, D. J. et al. Effective knockdown-replace gene therapy in a novel mouse model of DNM1 developmental and epileptic encephalopathy. Mol. Ther. 32, 3318–3330 (2024).

32. Bonnycastle, K. et al. Reversal of cell, circuit and seizure phenotypes in a mouse model of DNM1 epileptic encephalopathy. Nat. Commun. 14, 5285 (2023).

33. Sheffer, R. et al. Postnatal microcephaly and pain insensitivity due to a de novo heterozygous DNM1L mutation causing impaired mitochondrial fission and function. Am. J. Med. Genet. A. 170, 1603–1607 (2016).

34. Bartley, E. J. & Fillingim, R. B. Sex differences in pain: a brief review of clinical and experimental findings. Surv. Anesthesiol. 60, (2016).

35. Dance, A. Why the sexes don’t feel pain the same way. Nature 567, 448–450 (2019).

36. Arnold, A. P., Klein, S. L., McCarthy, M. M. & Mogil, J. S. Male–female comparisons are powerful in biomedical research — Don’t abandon them. Nature 629, 37–40 (2024).

37. Freidin, M. B. et al. Sex- and age-specific genetic analysis of chronic back pain. PAIN 162, (2021).

38. Dharshika, C. & Gulbransen, B. D. Enteric neuromics: how high-throughput “omics” deepens our understanding of enteric nervous system genetic architecture. Cell. Mol. Gastroenterol. Hepatol. 15, 487–504 (2023).

39. Delvalle, N. M. et al. Communication between enteric neurons, glia, and nociceptors underlies the effects of tachykinins on neuroinflammation. Cell. Mol. Gastroenterol. Hepatol. 6, 321–344 (2018).

40. Nemani, V. M. et al. Increased expression of α-synuclein reduces neurotransmitter release by inhibiting synaptic vesicle reclustering after endocytosis. Neuron 65, 66–79 (2010).

41. Wagh, D. et al. Piccolo directs activity dependent F-actin assembly from presynaptic active zones via Daam1. PLOS ONE 10, e0120093 (2015).

42. Pellegrino, A. et al. Differential expression of microRNAs in serum of patients with chronic painful polyneuropathy and healthy age-matched controls. Biomedicines 11, (2023).

43. Sun, Y.-Y. et al. TRIM32 deficiency impairs the generation of pyramidal neurons in developing cerebral cortex. Cells 11, (2022).

44. Sieghart, W. et al. Structure and subunit composition of GABAA receptors. Neurochem. Int. 34, 379–385 (1999).

45. Yang, S. & Chang, M. C. Chronic pain: structural and functional changes in brain structures and associated negative affective states. Int. J. Mol. Sci. 20, (2019).

46. Encinosa, W., Bernard, D. & Valdez, R. B. The association between smoking, chronic pain, and prescription opioid use: 2013-2021. J. Pain 26, (2025).

47. Johnston, K. J. A. & Huckins, L. M. Chronic pain and psychiatric conditions. Complex Psychiatry 9, 24–43 (2022).

48. Roughan, W. H. et al. Comorbid chronic pain and depression: shared risk factors and differential antidepressant effectiveness. Front. Psychiatry 12, (2021).

49. Zheng, C. J., Van Drunen, S. & Egorova-Brumley, N. Neural correlates of co-occurring pain and depression: an activation-likelihood estimation (ALE) meta-analysis and systematic review. Transl. Psychiatry 12, 196 (2022).

50. Husak, A. J. & Bair, M. J. Chronic pain and sleep disturbances: A pragmatic review of their relationships, comorbidities, and treatments. Pain Med. 21, 1142–1152 (2020).

51. Jain, S. V., Panjeton, G. D. & Martins, Y. C. Relationship between sleep disturbances and chronic pain: A narrative review. Clin. Pract. 14, 2650–2660 (2024).

52. Zhao, S. S., Holmes, M. V. & Alam, U. Disentangling the relationship between depression and chronic widespread pain: A Mendelian randomisation study. Semin. Arthritis Rheum. 60, 152188 (2023).

53. Tang, B., Meng, W., Hägg, S., Burgess, S. & Jiang, X. Reciprocal interaction between depression and pain: results from a comprehensive bidirectional Mendelian randomization study and functional annotation analysis. PAIN 163, (2022).

54. Yao, C. et al. Exploring the bidirectional relationship between pain and mental disorders: a comprehensive Mendelian randomization study. J. Headache Pain 24, 82 (2023).

55. Koller, D. et al. Pleiotropy and genetically inferred causality linking multisite chronic pain to substance use disorders. Mol. Psychiatry 29, 2021–2030 (2024).

56. Zhang, K. & Liang, H. Causal impacts of smoking on pain conditions and the mediating pathways: a mendelian randomization study. Sci. Rep. 14, 23375 (2024).

57. Grasby, K. L. et al. The genetic architecture of the human cerebral cortex. Science 367, eaay6690.

58. Morey, R. A. et al. Genomic structural equation modeling reveals latent phenotypes in the human cortex with distinct genetic architecture. Transl. Psychiatry 14, 451 (2024).

59. Farrell, S. F. et al. Genetic basis to structural grey matter associations with chronic pain. Brain 144, 3611–3622 (2021).

60. Neumann, N., Domin, M., Schmidt, C.-O. & Lotze, M. Chronic pain is associated with less grey matter volume in the anterior cingulum, anterior and posterior insula and hippocampus across three different chronic pain conditions. Eur. J. Pain 27, 1239–1248 (2023).

61. Farrell, S. F. et al. Genetic impact of blood C-reactive protein levels on chronic spinal & widespread pain. Eur. Spine J. 32, 2078–2085 (2023).

62. Morris, P., Ali, K., Merritt, M., Pelletier, J. & Macedo, L. G. A systematic review of the role of inflammatory biomarkers in acute, subacute and chronic non-specific low back pain. BMC Musculoskelet. Disord. 21, 142 (2020).

63. Kumbhare, D. et al. Potential role of blood biomarkers in patients with fibromyalgia: a systematic review with meta-analysis. PAIN 163, (2022).

64. Kasher, M. et al. Insights into the pleiotropic relationships between chronic back pain and inflammation-related musculoskeletal conditions: rheumatoid arthritis and osteoporotic abnormalities. PAIN 164, (2023).

65. Li, W. et al. Assessing the causal relationship between genetically determined inflammatory biomarkers and low back pain risk: a bidirectional two-sample Mendelian randomization study. Front. Immunol. 14, (2023).

66. Suri, P. et al. A Mendelian randomization study finds no evidence for causal effects of C-reactive protein on chronic pain conditions. Pain Med. pnae122 (2024) doi:10.1093/pm/pnae122.

67. Mills, S. E. E., Nicolson, K. P. & Smith, B. H. Chronic pain: a review of its epidemiology and associated factors in population-based studies. Br. J. Anaesth. 123, e273–e283 (2019).

68. Farrar, J. T. A consideration of differences in pain scales used in clinical trials. PAIN 163, (2022).

69. Euasobhon, P. et al. Reliability and responsivity of pain intensity scales in individuals with chronic pain. PAIN 163, (2022).

70. Willer, C. J., Li, Y. & Abecasis, G. R. METAL: fast and efficient meta-analysis of genomewide association scans. Bioinformatics 26, 2190–2191 (2010).

71. Watanabe, K., Taskesen, E., van Bochoven, A. & Posthuma, D. Functional mapping and annotation of genetic associations with FUMA. Nat. Commun. 8, 1826 (2017).

72. Auton, A. et al. A global reference for human genetic variation. Nature 526, 68–74 (2015).

73. Yang, J., Lee, S. H., Goddard, M. E. & Visscher, P. M. GCTA: A tool for genome-wide complex trait analysis. Am. J. Hum. Genet. 88, 76–82 (2011).

74. Benner, C. et al. FINEMAP: efficient variable selection using summary data from genome-wide association studies. Bioinformatics 32, 1493–1501 (2016).

75. de Leeuw, C. A., Mooij, J. M., Heskes, T. & Posthuma, D. MAGMA: Generalized gene-set analysis of GWAS data. PLOS Comput. Biol. 11, e1004219 (2015).

76. Guo, Y. et al. SMR-Portal: an online platform for integrative analysis of GWAS and xQTL data to identify complex trait genes. Nat. Methods (2024) doi:10.1038/s41592-024-02561-7.

77. Sey, N. Y. A. et al. A computational tool (H-MAGMA) for improved prediction of brain-disorder risk genes by incorporating brain chromatin interaction profiles. Nat. Neurosci. 23, 583–593 (2020).

78. Qi, T. et al. Genetic control of RNA splicing and its distinct role in complex trait variation. Nat. Genet. 54, 1355–1363 (2022).

79. The GTEx Consortium et al. The GTEx Consortium atlas of genetic regulatory effects across human tissues. Science 369, 1318–1330 (2020).

80. McRae, A. F. et al. Identification of 55,000 replicated DNA methylation QTL. Sci. Rep. 8, 17605 (2018).

81. Zheng, J. et al. Phenome-wide Mendelian randomization mapping the influence of the plasma proteome on complex diseases. Nat. Genet. 52, 1122–1131 (2020).

82. Sun, B. B. et al. Plasma proteomic associations with genetics and health in the UK Biobank. Nature 622, 329–338 (2023).

83. Wingo, T. S. et al. Shared mechanisms across the major psychiatric and neurodegenerative diseases. Nat. Commun. 13, 4314 (2022).

84. Zhu, Z. et al. Integration of summary data from GWAS and eQTL studies predicts complex trait gene targets. Nat. Genet. 48, 481–487 (2016).

85. Grotzinger, A. D. et al. Genomic structural equation modelling provides insights into the multivariate genetic architecture of complex traits. *Nat*. Hum. Behav. 3, 513–525 (2019).

86. Hemani, G. et al. The MR-Base platform supports systematic causal inference across the human phenome. eLife 7, e34408 (2018).

87. Burgess, S., Thompson, S. G., & CRP CHD Genetics Collaboration. Avoiding bias from weak instruments in Mendelian randomization studies. Int. J. Epidemiol. 40, 755–764 (2011).

88. Palmer, T. M. et al. Using multiple genetic variants as instrumental variables for modifiable risk factors. Stat. Methods Med. Res. 21, 223–242 (2012).

89. Hemani, G., Tilling, K. & Davey Smith, G. Orienting the causal relationship between imprecisely measured traits using GWAS summary data. PLOS Genet. 13, e1007081 (2017).

90. Lin, S. et al. Inferring the genetic relationship between brain imaging-derived phenotypes and risk of complex diseases by Mendelian randomization and genome-wide colocalization. NeuroImage 279, 120325 (2023).

91. Frei, O. et al. Bivariate causal mixture model quantifies polygenic overlap between complex traits beyond genetic correlation. Nat. Commun. 10, 2417 (2019).

92. Holland, D. et al. Beyond SNP heritability: Polygenicity and discoverability of phenotypes estimated with a univariate Gaussian mixture model. PLOS Genet. 16, e1008612 (2020).

93. Said, S. et al. Genetic analysis of over half a million people characterises C-reactive protein loci. Nat. Commun. 13, 2198 (2022).

94. Als, T. D. et al. Depression pathophysiology, risk prediction of recurrence and comorbid psychiatric disorders using genome-wide analyses. Nat. Med. 29, 1832–1844 (2023).

95. Nagel, M. et al. Meta-analysis of genome-wide association studies for neuroticism in 449,484 individuals identifies novel genetic loci and pathways. Nat. Genet. 50, 920–927 (2018).

96. Wu, Y. et al. Genome-wide association study of medication-use and associated disease in the UK Biobank. Nat. Commun. 10, 1891 (2019).

97. Nievergelt, C. M. et al. Genome-wide association analyses identify 95 risk loci and provide insights into the neurobiology of post-traumatic stress disorder. Nat. Genet. 56, 792–808 (2024).

98. Khan, Y. et al. Combining transdiagnostic and disorder-level GWAS enhances precision of psychiatric genetic risk profiles in a multi-ancestry sample. medRxiv 2024.05.09.24307111 (2024) doi:10.1101/2024.05.09.24307111.

99. Roden, D. et al. Development of a large-scale de-identified DNA biobank to enable personalized medicine. Clin. Pharmacol. Ther. 84, 362–369 (2008).

100. Purcell, S. et al. PLINK: A tool set for whole-genome association and population-based linkage analyses. Am. J. Hum. Genet. 81, 559–575 (2007).

101. McCarthy, S. et al. A reference panel of 64,976 haplotypes for genotype imputation. Nat. Genet. 48, 1279–1283 (2016).

102. Fuchsberger, C., Abecasis, G. R. & Hinds, D. A. minimac2: faster genotype imputation. Bioinformatics 31, 782–784 (2015).

103. Das, S. et al. Next-generation genotype imputation service and methods. Nat. Genet. 48, 1284–1287 (2016).

104. Ge, T., Chen, C.-Y., Ni, Y., Feng, Y.-C. A. & Smoller, J. W. Polygenic prediction via Bayesian regression and continuous shrinkage priors. Nat. Commun. 10, 1776 (2019).

105. Denny, J. C., Bastarache, L. & Roden, D. M. Phenome-wide association studies as a tool to advance precision medicine. Annual Review of Genomics and Human Genetics vol. 17 353–373 (2016).

